# Is age the most important risk factor in COVID-19 patients? The relevance of comorbidity burden: A retrospective analysis of 10,090 hospitalizations

**DOI:** 10.1101/2022.06.14.22276380

**Authors:** Damià Valero-Bover, David Monterde, Gerard Carot-Sans, Miguel Cainzos-Achirica, Josep Comin-Colet, Emili Vela, Montse Clèries, Sònia Abilleira, Miquel Arrufat, Yolanda Lejardi, Òscar Solans, Toni Dedeu, Marc Coca, Pol Pérez-Sust, Jordi Piera-Jiménez

**Affiliations:** Catalan Health Service, Barcelona (Spain); Digitalization for the Sustainability of the Healthcare System (DS3) – Institut d’Investigacions Biomèdiques de Bellvitge (IDIBELL), Barcelona (Spain); Catalan Institute of Health, Barcelona (Spain); Center for Outcomes Research, Houston Methodist, Houston, TX (USA); Johns Hopkins Ciccarone Center for the Prevention of Cardiovascular Disease, Johns Hopkins Medical Institutions, Baltimore, MD, (USA); Cardiology Department, Bellvitge University Hospital (IDIBELL), Barcelona (Spain); University of Barcelona, Hospitalet de Llobregat, Barcelona (Spain); CIBER Epidemiología y Salud Pública (CIBERESP), Barcelona (Spain); Health Department, eHealth Unit, Barcelona (Spain); WHO European Centre for Primary Health Care, Almaty (Kazakhstan); Faculty of Informatics, Telecommunications and Multimedia, Universitat Oberta de Catalunya, Barcelona (Spain)

## Abstract

**Objectives:** To investigate whether the effect of chronological age on the risk of developing critical illness in COVID-19 hospitalized individuals is attenuated, and to which extent, when adjusting for increasingly exhaustive measures of the comorbidity burden.

**Design:** Retrospective assessment of electronic health records.

**Setting:** All public hospitals of the Catalan Institute of Health (Catalonia; North-East Spain; 7.7 million inhabitants), which account for 30% of all hospital admissions in Catalonia.

**Participants:** We included all individuals admitted to the hospital with COVID-19 as the main diagnosis between March 1, 2020, and January 31, 2022. Vaccinated individuals and those admitted within the first of the six COVID-19 epidemic waves were excluded from the primary analysis but were included in secondary analyses.

**Main outcome measures:** The primary composite outcome was critical illness, defined as the need for invasive mechanical ventilation, transfer to the intensive care unit (ICU), or in-hospital death (any of them). Explanatory variables included age, sex, and four summary measures of comorbidity burden on admission: the Charlson index (17 diagnostic group codes), the Elixhauser index and count (31 diagnostic group codes), and the Queralt DxS index (3,145 diagnostic group codes). All models were adjusted by wave and center. The proportion of the effect of age attributable to comorbidity burden was assessed using a causal mediation analysis.

**Results:** The primary analysis included 10,090 hospitalizations due to COVID-19; of them, 3,524 experienced a critical illness. The frequency of critical illness increased with age and comorbidity burden on admission, irrespective of the measure used. In multivariate analyses, the effect size of age decreased with the number of diagnoses considered to estimate comorbidity burden. When adjusting for the Queralt DxS index, age showed a minimal contribution to critical illness; according to the causal mediation analysis, comorbidity burden on admission explained the 95.3% (95% CI 82.1% −112.7%) of the observed effect of age on critical illness.

**Conclusions:** When measured exhaustively, comorbidity burden rather than chronological age explains the increased risk of critical illness observed in patients hospitalized with COVID-19.

**Summary box:** *What is already known on this topic:*

- Age is broadly acknowledged as a critical risk factor for developing critical illness in individuals hospitalized due to COVID-19.
- When adjusting for other underlying factors, such as comorbidities, the effect size of age for predicting critical illness decreases; nevertheless, most studies have suggested that age remains independently associated with COVID-19 outcomes.

*What this study adds:*

- The observed contribution of chronological age to the risk of critical illness in hospitalized COVID-19 patients decreases with the exhaustivity of the measure of comorbidity burden.
- When adjusting for a comprehensive comorbidity index that considers all possible clinical conditions from a weighted list of 3,145 possible diagnostic groups, age has little or no relevant effect on the risk of critical illness.
- Mediation analyses confirm that the effect of chronological age on COVID-19 outcomes can be explained by comorbidity burden.

## Introduction

Since the beginning of the coronavirus disease (COVID-19) pandemic, age and the presence of comorbidities have both been pointed out as critical risks factors for developing severe illness [1]. Various authors have found that the influence of age on severe outcomes decreased when adjusting for other factors, including but not limited to comorbidities [2–4]. Nevertheless, in most models such attenuation of the effect of age was only partial, and age was still acknowledged as the most important risk factor for severe illness [5–7]. In this context, age has been used as the main criterion for prioritizing vaccine allocation in many countries and driving many stratify-and-shield campaigns worldwide.

While the effect of age can be easily measured in multivariate models, assessing the contribution of comorbidity burden has several challenges, which may bias the results. Most studies in the COVID-19 setting have assessed the presence or absence of a specific, relatively limited number of chronic conditions [5,6,8–11]. However, this approach addresses the effect of certain comorbidities rather than the effect of comorbidity burden as a whole. Alternatively, other authors have used summary measures of comorbidity burden, such number of chronic conditions (e.g., stratified into categories from 0 to up to ≥ 3) [6,12,13], or summary indices such as the Charlson or Elixhauser [14–19]. These indices might underestimate comorbidity burden due to the limited number of diagnoses considered [20].

Taking advantage of the systematic collection and integration of routine care data, we recently developed a set of comprehensive indices for risk assessment in hospitalized patients, which includes an index for measuring comorbidity burden of these patients: the Queralt index for comorbidities (Queralt DxS) [21]. The index combines and weighs more than 3,145 relevant acute and chronic diagnostic codes and provides a numerical index of comorbidity burden on admission. When used as adjustment factor in risk assessment of patients hospitalized with COVID-19, the Queralt DxS showed a remarkable contribution to explaining the risk of critical illness (i.e., admission to intensive care unit or death) in individuals hospitalized with COVID-19 [22].

In this analysis, we investigated how the effect of chronological age on the risk of critical illness changes when comorbidity burden is measured using increasingly exhaustive tools: the Charlson index (17 diagnostic group codes), the Elixhaurser index (31 diagnostic group codes), the count of diagnoses included in the Elixhauser index, and the Queralt DxS (3,145 diagnostic group codes). We also aimed at evaluating the mediation role of comorbidity burden in the relationship between age and critical COVID-19 illness.

## Methods

### Study design and setting

This was a retrospective analysis of individuals hospitalized with COVID-19 as the primary diagnosis in the seven public hospitals of the Catalan Institute of Health, the leading healthcare provider in Catalonia (North-East Spain). The Catalan Institute of Health provides universal healthcare to nearly 70% of the Catalan population and accounts for 30% of the total acute hospitalizations in Catalonia.

We screened the database of the Catalan Institute of Health for all individuals admitted with COVID-19 as the primary diagnosis between March 1, 2020 and January 31, 2022. Data were extracted on May 10, 2022. Patients derived from other hospitals or transferred to other hospitals on discharge were excluded from the record. For the primary diagnosis, we considered the following diagnosis codes of the international classification of diseases 10^th^ version, clinical modification (ICD-10-CM): B97.29, B97.21, B34.2, J12.81, J12.89, and U07.1. The vaccination campaign in Catalonia started on December 27, 2020. Figure S1 (Supplementary file 1) summarizes the prevalence of each variant of concern throughout the investigated period.

All data were handled according to the General Data Protection Regulation 2016/679 on data protection and privacy for all individuals within the European Union and the local regulatory framework regarding data protection. The study protocol was approved by the independent ethics committee of the Bellvitge Biomedical Research Institute (IDIBELL, Spain), which waived obtaining informed consent for the secondary use of data collected during routine care (ref. PR123/22).

### Variables and data sources

The study outcome was a composite outcome of critical illness, which included the need for invasive mechanical ventilation, transfer to the intensive care unit (ICU), or in-hospital death (any of them). Information about admission to ICU and death are systematically collected in the electronic health records of Catalan Institute of Health hospitals, whereas the need for invasive mechanical ventilation was determined by the following codes of hospital procedures: 5A09357, 5A09457, 5A09557, 5A1935Z, 5A1945Z, 5A1955Z, 09HN7BZ, 09HN8BZ, 0BH13EZ, 0BH17EZ, 0BH18EZ, 0CHY7BZ.

Primary explanatory variables included age, sex, and measures of the comorbidity burden present on admission. We used four summary measures of comorbidity burden: the Charlson index [23], the Elixhauser count (i.e., number of diagnoses among the 31 codes considered in the Elixhauser index) and index [24], and the Queralt index for secondary diagnoses present on admission (Queralt DxS) [21]. The ICD-10 coding for the Charlson and Elixhauser scores was based on work by Quan et al. [25]. Weights for the Charlson score are based on the original formulation by Charlson et al. in 1987 [23], while weights for the Elixhauser score were based on work by Moore et al. [26]. The Queralt DxS is part of a set of three indices for measuring the clinical complexity of hospitalized patients. It provides a numerical value from the weighted sum of secondary diagnoses present on admission from a list of 3,145 diagnostic code groups [21]. The weights of the version used in this analysis (version 6.3) were estimated from health data collected in hospitalizations reported between 2018 and 2019 in the Catalan Institute of Health and were, therefore, not specific to COVID-19 patients.

Adjusting variables included the wave in which the admission occurred, the hospital, and the vaccination status for COVID-19 (the last used only in the secondary analyses presented in the supplementary material). The vaccination status was retrieved from the K2 platform database, held by the Catalan Department of Health and used as a source of information for issuing COVID-19 certificates. The K2 database includes information on COVID-19 diagnoses from the primary and hospital care setting and vaccination information. Vaccination categories, used in the secondary analysis only, were defined as follows: partial vaccination (i.e., one dose of a 2-dose regimen of an RNA-based vaccine), complete vaccination (i.e., either two doses of an RNA vaccine or one dose of a single-dose regimen vaccine), and booster (i.e., an additional dose to the complete vaccination regimen).

### Analysis

For the primary analysis, we excluded hospitalizations that occurred during the first wave of the COVID-19 outbreak in Catalonia (from March 1 to June 23, 2020). We expected those to be associated with a significant risk of bias, as the initial outbreak in Spain overwhelmed hospital resources, knowledge on the management of COVID-19 in the hospital setting was very limited [11,27], and data collection in this setting was of limited quality. We also excluded individuals who received at least one vaccine dose from the primary analysis. Secondary analyses included the whole cohort of patients hospitalized with COVID-19 as the primary diagnosis (irrespective of the vaccine status) within the entire period, stratified according to waves.

For description purposes, age and the summary measures of comorbidity were categorized. Age was split into the following groups: 0 - 39, 40 - 49, 50 - 59, 60 - 69, 70 - 79, and ≥ 80). The indices of comorbidity were categorized into three risk levels of comorbidity burden that yield homogeneously sized high-risk groups: the Charlson index scores were grouped into low health risk (score 0), moderate (1 – 2), and high (≥ 3); the Elixhauser and Queralt DxS indices were grouped into low (below the 50^th^ percentile), moderate (50^th^ – 85^th^), and high (>85^th^ percentile); the Elixhauser count was grouped into low (0 – 1), moderate (2 – 3), and high (≥ 4). The definition of the cut-off percentiles for the Elixhauser and Queralt DxS indices sough homoscedasticity with age (i.e., 15% of the study sample was allocated in the upper age group).

The association between explanatory variables (age, sex, and comorbidity burden) and the study outcome (development of critical illness) was investigated using multiple logistic regression models for each measure of the comorbidity burden: Charlson index, Elixhauser index, Elixhauser count, and Queralt DxS index. Age was introduced as a categorical variable to ease the interpretation of the resulting model, although the same models with age as a continuous variable plus an additional quadratic term were built to confirm the equivalent performance of the model. In addition, and acknowledging potential clustering of patient characteristics by hospital [27], all models were further adjusted by considering the random effects of hospitals in which admission occurred. First, we built separate models for age, sex, and each measure of the comorbidity burden; then, we built multivariate models including age, sex, and one comorbidity measure; finally, we built the same models accounting for interactions between age and the comorbidity measures. The same methodology was applied to secondary analyses in which each wave was analyzed separately, with models adjusted for hospital and vaccination status.

Finally, we used a causal mediation analysis [28] to investigate the hypothesis that comorbidity burden, would fully mediate the association between age (exposure factor or treatment variable) and critical illness (outcome). In this analysis, age and the comorbidity indices were used as continuous variables. The control and treatment age groups, required for the mediation analysis, were established based on the 50 and 75 years cutoffs. The average causal mediation effect of comorbidity (mediator), the average direct effect of age, and the total effect were estimated, and the 95% CI obtained using bootstrap from 2,000 simulations, considered adequate for this type of analysis. The contribution of the comorbidity-mediating pathway was assessed using the proportion of the average causal mediation effect over the total effect. All analyses were performed using R 4.1.2 [29]. The causal mediation analysis was conducted using the library mediation by Tingley et al. [30], the linear and mixed model adjustments were conducted using the lme4 library [31], and the analyses of the ROC and precision-recall curves were done using the pROC [32] and PRROC [33] libraries, respectively. The Charlson and Elixhauser indices were computed using the Comorbidity library by Gasparini [34], whereas the Queralt DxS was estimated using the updated version of the index R function (version 6.3, which are available from the corresponding author for research purposes.

## Results

### Study population

Between March 1, 2020 and January 31, 2022, 15,717 individuals were admitted to the hospitals of the Catalan Institute of Health with COVID-19 as the main diagnosis (Figure 1). Of them, 10,090 were non-vaccinated individuals admitted after the first wave (i.e., from June 23, 2020 on) and were, therefore, included in the primary analysis: 3,524 (35%) with critical illness and 6,566 (65%) without critical illness. The second wave contributed the largest number of admissions to this analysis.

**Figure 1.**
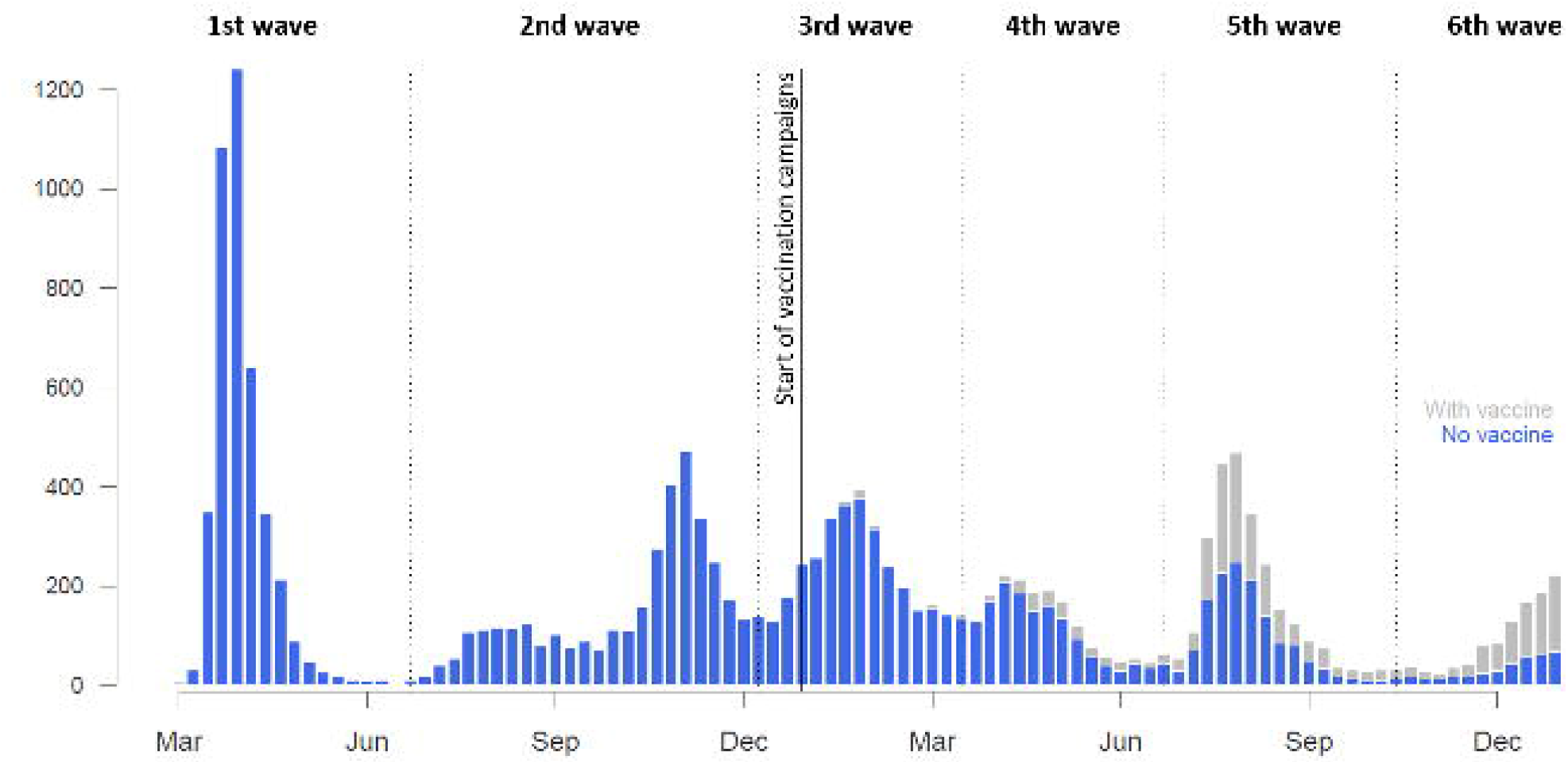
Absolute number of weekly admissions throughout the period. Individuals in the no vaccine group had not received any dose of any type of vaccine against COVID-19. Vertical dotted lines indicate the start/end of each COVID-19 wave in our area.

Table 1 summarizes the main demographic, epidemiological, and clinical characteristics of the study population of the primary analysis. The characteristics according to COVID-19 wave are listed in Tables S1 to S6. The proportion of patients with critical illness increased with age and comorbidity burden, irrespective of the type of measure used. The same trends were observed when age and comorbidity burden were described as continuous variables. The greatest differences in the proportion of critical illness according to comorbidity were observed for the Queralt DxS.

**Table 1.**
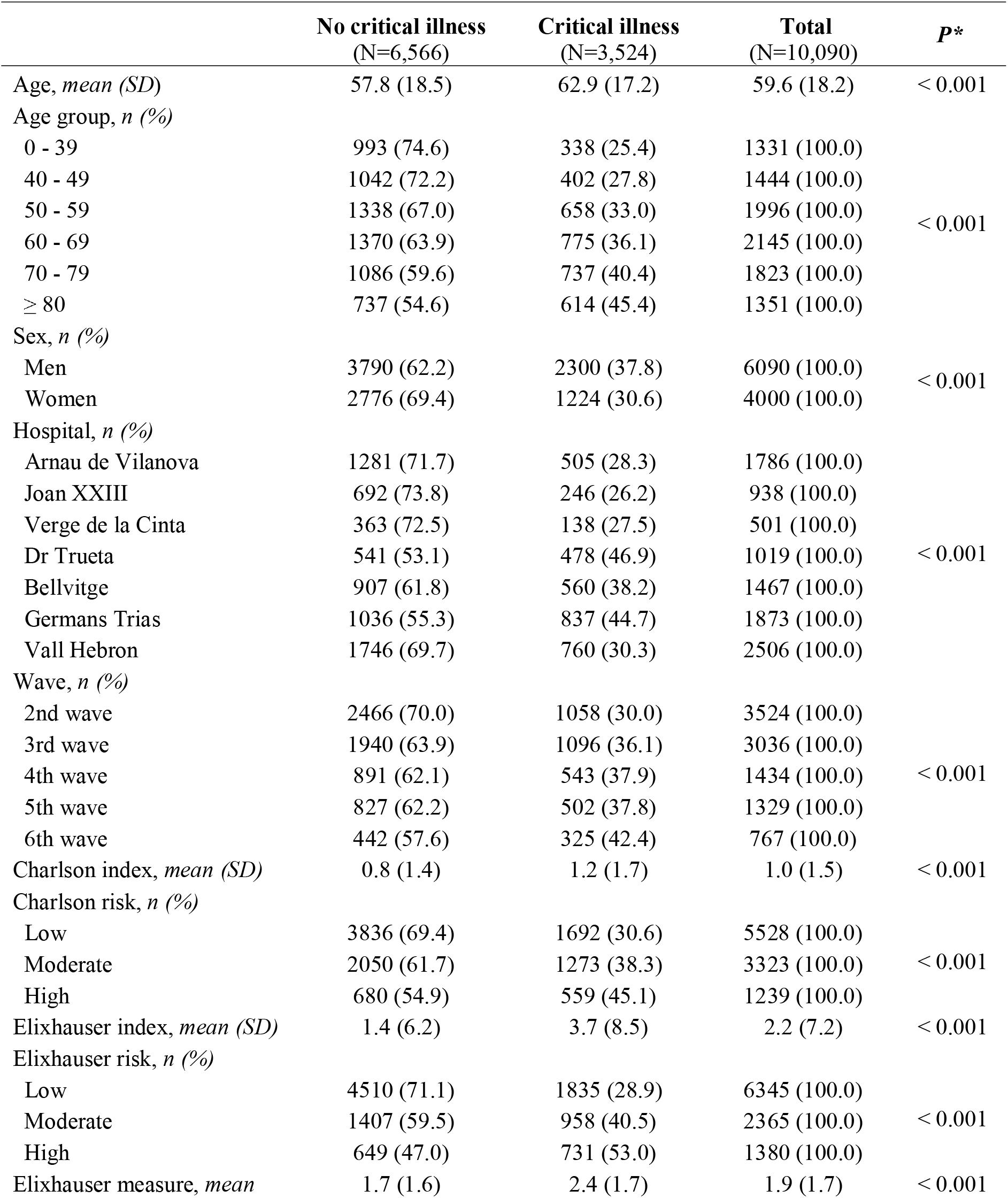
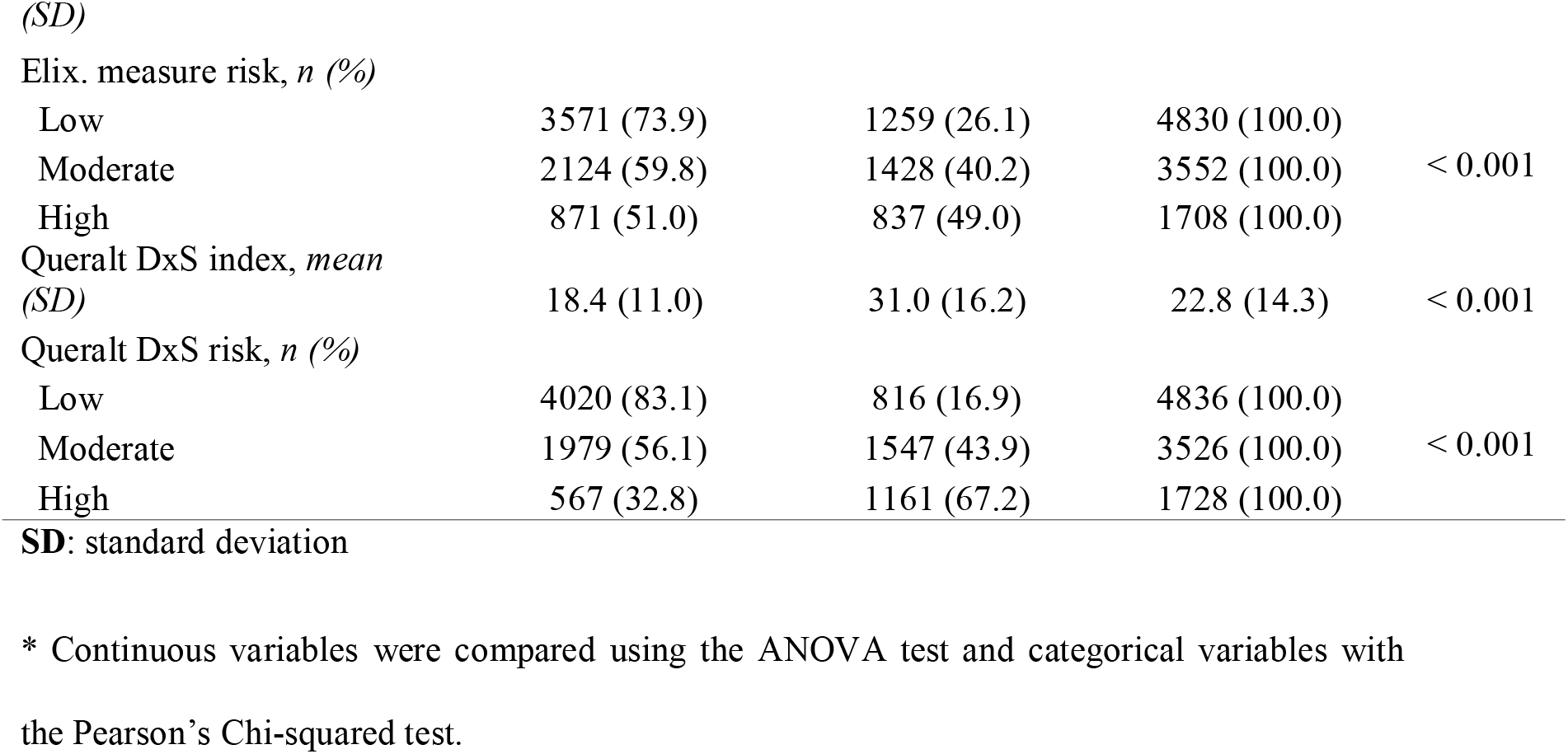
Demographic, clinical, and epidemiological characteristics of individuals included in the primary analysis.

### Estimated risk and critical illness

The distribution of the study population across the Queralt DxS risk groups showed a higher proportion of individuals at high and moderate risk among patients who experienced critical illness (Figure 2). The distribution according to the successive waves showed a similar trend (Figures S2-S7).

**Figure 2.**
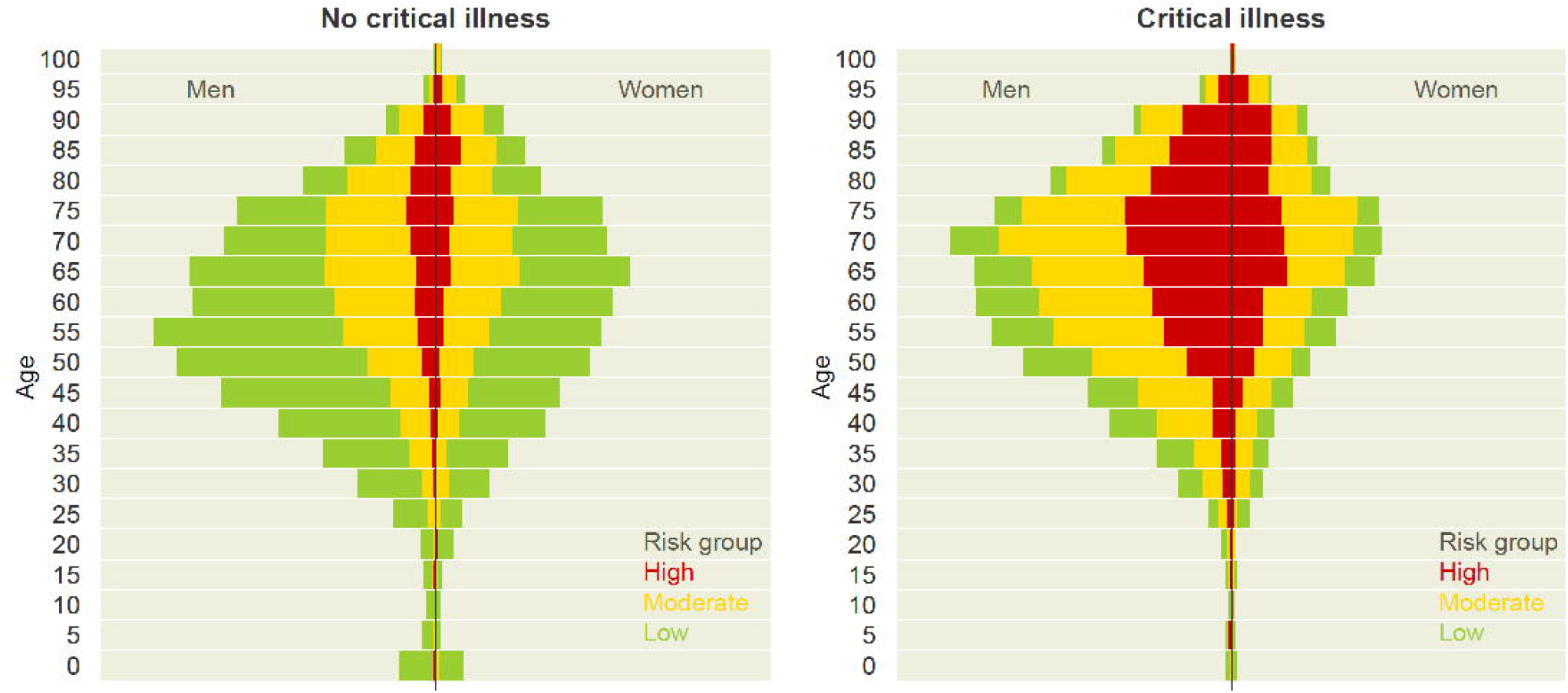
Distribution of Queralt DxS-based risk groups, by age and sex, among individuals who did not and did develop critical illness. Primary analysis population (N=10,090)

### Risk factors for critical illness

The bivariate analyses showed that age and comorbidity burden, irrespective of the index used, significantly increased the risk of critical illness (Figure S8). Moreover, the risk of critical illness increased linearly with both factors. Older age showed a strong effect on the risk of death, whereas comorbidity burden measured with the Queralt DxS showed the largest effect on the composite outcome of critical illness. This observation was consistent across the successive waves (Figures S9-S14).

According to the baseline model, adjusted by age, sex, hospital, and wave, the risk of critical illness increased linearly with age and was higher in men (Figure 3). When adjusting also for summary indices of comorbidity, the effect size of sex remained relatively stable, whereas that of age progressively decreased with the exhaustivity of the comorbidity measure. The reduction of the effect size of age was the highest when comorbidity burden was summarized using the Queralt DxS (i.e., the comorbidity measure that considers the highest number of possible diagnostic groups). We observed the same trend in all waves, except the first one, in which the effect of age remained significant for all age groups above 60 years when adjusting for the Queralt DxS (Figure S15 to S20).

**Figure 3.**
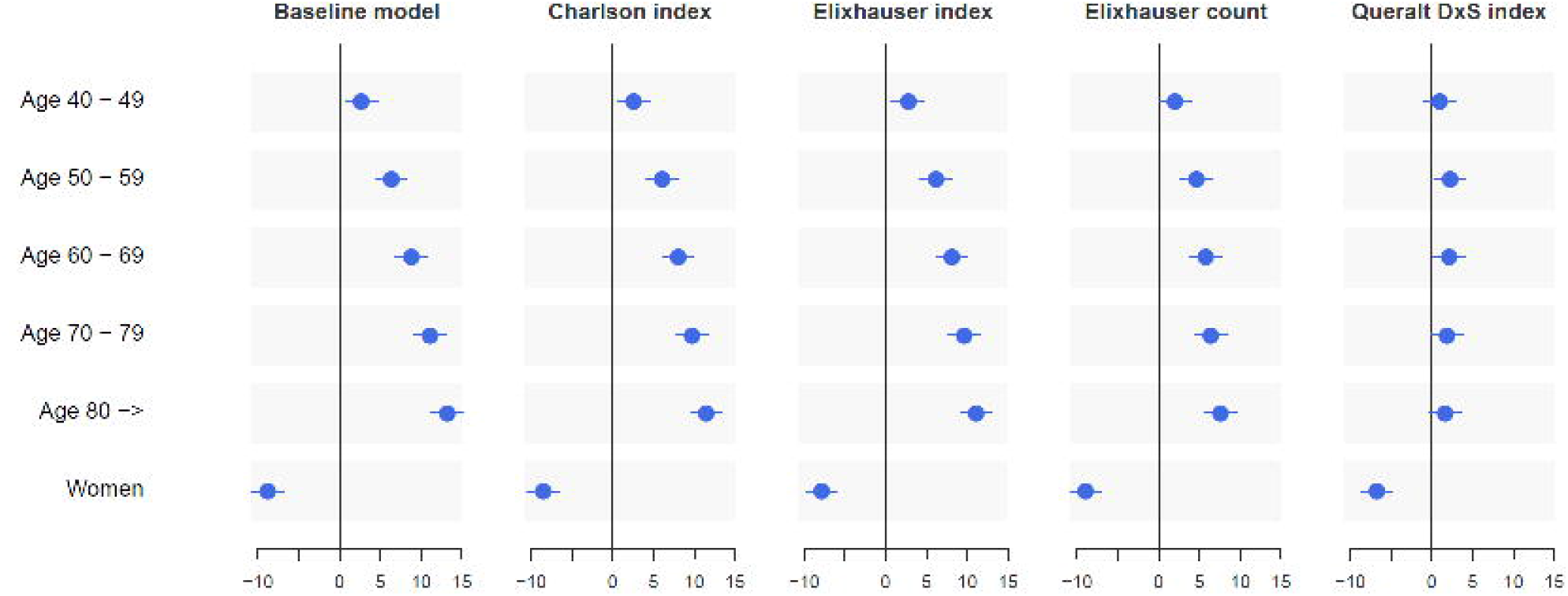
Effect of age (standardized coefficients for age groups) and sex on critical illness using multiple logistic regressions. The baseline model included only age, sex, and hospital, whereas the other models were built by adjusting the baseline model for each of the multimorbidity measures: the Charlson index, the Elixhauser index, the unweighted count of Elixhauser diagnoses, and the Queralt DxS.

### Performance of prediction models for critical illness

Table 2 summarizes the BIC, AUROC, and precision-recall estimates for models in three series of models: (1) models including only sex, age, or a summary measure of the comorbidity burden (adjusted by hospital and wave), (2) multivariate models including age, sex, and a comorbidity measure (also adjusted by hospital and wave), and (3) the corresponding models accounting for interactions between age and the comorbidity measures. In all model series, the performance increased with the number of diagnoses considered for the comorbidity burden estimate, with models using the Charlson index showing the poorest performance and models using the Queralt DxS the highest. The corresponding analyses for each wave are shown in Tables S7 to S12.

**Table 2.**
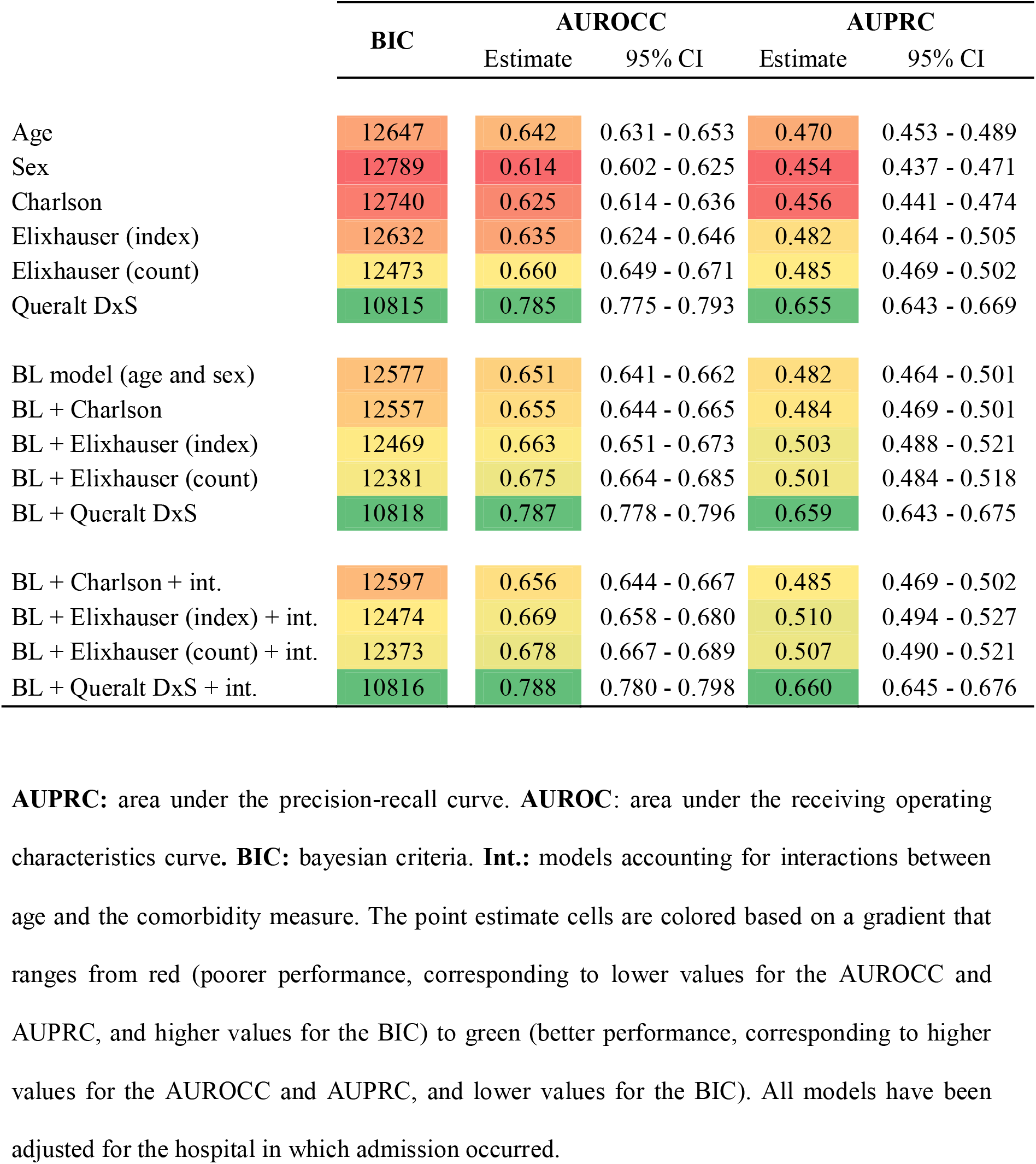
Performance of the models for explaining critical illness.

### Mediation analyses

According to the causal mediation analysis, the proportion of mediation by comorbidity burden over the total effect of age on the risk of critical illness increased with the exhaustivity of the comorbidity measure used (Figure 4). Using the Charlson index (i.e., the comorbidity measure with the smallest effect size in our multivariate analysis), the proportion of the effect that was mediated was remarkably lower than the direct effect of age. In contrast, the reverse was true when comorbidity burden was measured using the Queralt DxS. In the latter analysis, the direct effect of age was no longer significant (Figure 4, Panel D). The proportion of mediation effects was highest for the Queralt DxS index in all waves of the COVID-19 outbreak in Catalonia, with values ranging from 0.68 (95% CI 0.59-0.79) to 1.17 (95% CI 0.89-1.65) (Tables S13-S18).

**Figure 4.**
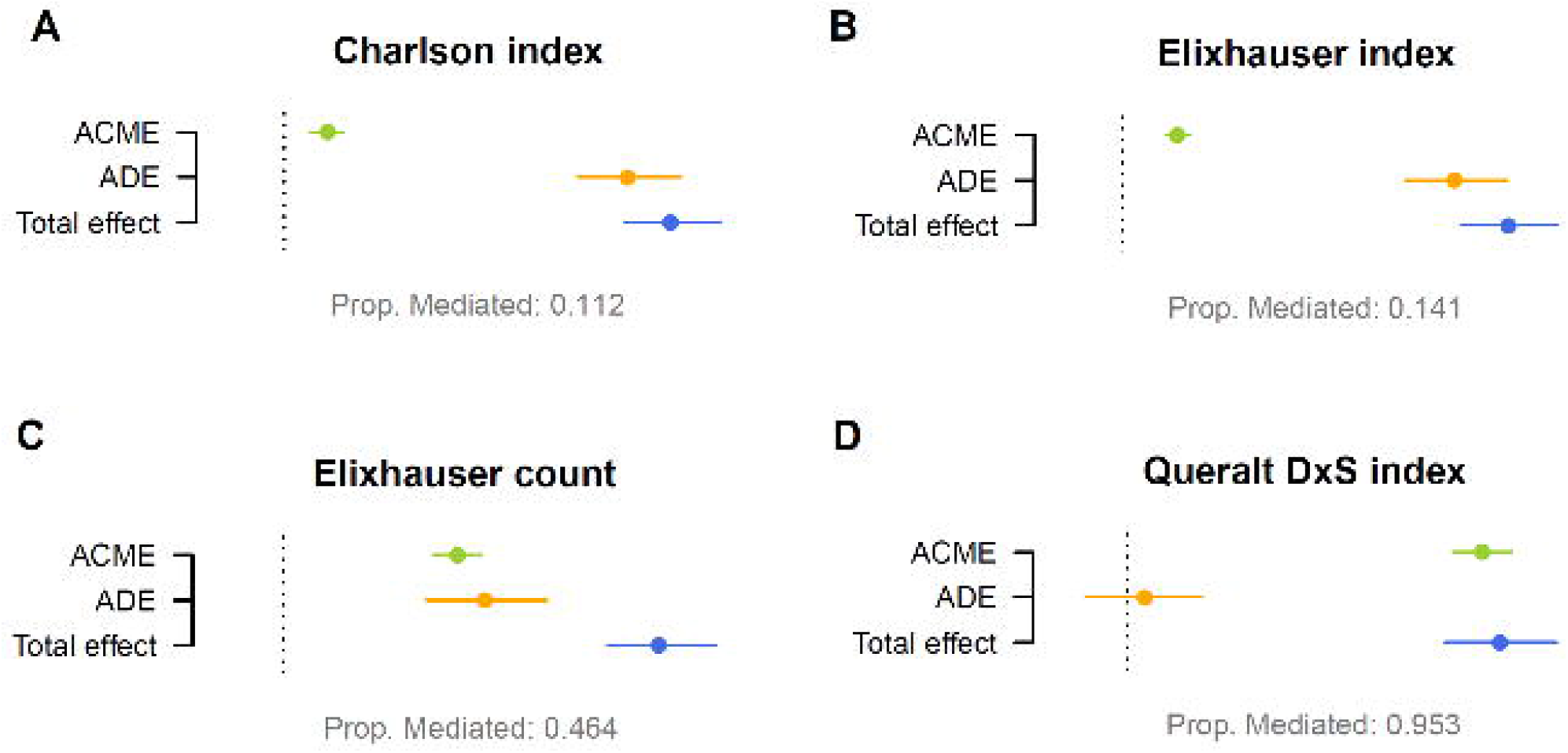
Causal mediation analysis: role of comorbidity burden on the association between age and incidence of critical illness, with comorbidity burden assessed using the Charlson index (**A**), the Elixhauser index (**B**), the Elixhauser count (**C**), and the Queralt DxS index (**D**). The proportion mediated shows the contribution of the comorbidity-mediating pathway to critical illness, estimated as the proportion between the ACME and the total effect. **ACME**: average causal mediation effect of comorbidity (mediator). **ADE**: average direct effect of age. The dotted line indicates the null value of the effect.

## Discussion

### Principal findings

In this retrospective analysis of 15,717 hospitalizations due to COVID-19, we found that the contribution of comorbidity burden to critical illness increases with the number of diagnoses considered for its measurement. When measured using a comprehensive index such as the Queralt DxS, which considers more than 3,000 possible diagnoses, the contribution of age to critical illness was remarkably reduced, with age groups 70 – 79 years and >80 years no longer associated with the odds of developing critical illness, and comorbidity burden explaining a significant proportion of the effect of chronological age on this outcome.

### Strengths and weaknesses of the study

Our analysis is strengthened by the exhaustive data collection of all diagnoses routinely reported in the hospital records in our region. This feature, along with the provision of universal public healthcare to all residents in Catalonia, allowed us to identify and consider in our models all concomitant clinical conditions in patients admitted to the hospital because of COVID-19. On the other hand, the use of administrative databases of routine care data has some limitations that must be considered for result interpretation. The most important drawback is the constraint of the analysis to the information recorded in the databases. Thus, aside from the diagnoses, other clinical conditions not recorded in these databases may play a role in the observed effect of age. These conditions include —but are not limited to— frailty (i.e., physical deterioration, not considered a diagnosis per se), weight loss, mild cognitive decline (i.e., not qualifying for dementia), the recent loss of a relative, or subclinical depression. These conditions are likely to impact mortality, regardless of the presence of comorbidities [35,36]. Therefore, they should also be considered to understand the effect of chronological age on COVID-19 outcomes completely. Nevertheless, the features captured by Queralt DxS were already able to explain almost the full effect of age on the odds of developing critical COVID-19 disease.

### Context with other studies

The question of whether age is the most important factor in explaining COVID-19 outcomes has been addressed previously. Semenzato et al. concluded that age was the most important factor based on the individual hazards of an extensive list of 47 comorbidities [5]. However, the authors acknowledged that the sum of the number of comorbidities does not account for the different severity of each of them. Henkens et al. followed a similar approach using a shorter list of comorbidities [6]. Additionally, the authors investigated the mediation effect of each comorbidity on the effect of age on in-hospital mortality and found that the direct effect of age was ≥95% in all diagnoses. However, when adjusting for the comorbidity burden, they used an unweighted count of diagnoses, stratified as 0, 1-2, and >2 comorbidities. These two approaches (i.e., the risk estimate of each diagnosis independently and the unweighted count of the number of diseases) lose sight of the actual disease burden and are likely to underestimate the effect of comorbidity burden as a whole, particularly in patients with relevant diagnoses simultaneously.

Alternatively, we measured the comorbidity burden using a very comprehensive index that considers more than 3,000 possible diagnoses and weights them according to their impact on health outcomes [21]. This measure has shown a high capacity for explaining hospital outcomes in other settings. Our results regarding the explaining capacity of this variable for COVID-19 outcomes were consistent across various waves of the outbreak in our region and in different analysis approaches. This trend was weaker in patients admitted during the first wave, which were excluded from the primary analysis. The extreme demand for ICU beds during the first wave exceeded by far the ICU capacity in our region. In this context, the criteria for ICU admission were unclear, and we cannot rule out an age bias in ICU transfers. This limitation was also noted by Henkens et al. [6]. Furthermore, retrospective analyses of the clinical presentation and hospital outcomes throughout successive waves have highlighted the progressive consolidation of evidence-based practices in the management of COVID-19 patients in the hospital setting [11,27], which may contribute to explaining the differences between waves. Moreover, during the first wave, quick decision making in a context of overwhelmed systems and a very high number of cases might have resulted in suboptimal documentation of chronic comorbidities.

### Implications for clinicians and policymakers

Our findings have various implications. First, in light of the prognostic importance of comorbidity burden as a whole, comprehensive and weighted metrics of this variable may increase accuracy of risk estimate, as suggested by Semenzano et al. [5]. If all diagnoses are adequately reported in the electronic health records, the Quearlt DxS, freely available for research purposes, might be used to summarize the comorbidity burden in a single index used as an adjustment covariate. The same adjustment can be used to develop predictive models using machine learning approaches, in which the summary of multiple comorbidities into a single weighted index may prevent overfitting. These models would allow identifying patients at higher risk of critical illness and creating stratification systems for prioritizing and allocating healthcare resources such as COVID-19 vaccines or anticipating the demand of hospital services. Finally, in healthcare systems that integrate primary and hospital care data [37], comorbidity burden could be estimated in advance to anticipate resource prioritization.

### Conclusions and future work

Our findings highlight the importance of considering the comorbidity burden as a whole (rather than individual diagnoses) and in a comprehensive way for assessing the risk of critical illness in COVID-19 patients. This perspective encourages the digitalization of healthcare systems for the systematic collection and integration of healthcare data that provides an accurate view of the clinical complexity of patients. Future steps in this pathway include the external validation of this tool and the inclusion of social care and functional information in these records.

In summary, although age is often regarded a key prognostic factor in people hospitalized with COVID-19, our study suggests that when measured exhaustively, comorbidity burden rather than chronological age explains the higher risk of critical illness observed as age increases.

Moving forward, greater attention to comorbidity burden rather than to chronological age may inform more accurate risk stratification, management, and preventive therapy allocation.

## Data Availability

All data produced in the present study are available upon reasonable request to the authors

## Declarations

### Ethics approval

The study protocol was approved by the independent ethics committee of the Bellvitge Biomedical Research Institute (IDIBELL, Spain), which waived obtaining informed consent for the secondary use of data collected during routine care (ref. PR123/22).

### Transparency

DM declares that he is the developer of the Queralt System. This tool is available online for research purposes at no cost. The authors report no other conflicts of interest in this work. All authors met the authorship criteria of the International Committee of Medical Journal Editors and were, therefore, listed as authors in the manuscript. DM had full access to the database used in this analysis.

## Acknowledgement

The authors would like to thank Toni Fuentes for his support in building the dataset used in this analysis.

## Patient and public involvement

In this observational, retrospective study, we used a full-cohort approach to the analysis, which involved 15,717 people. Therefore, patient involvement in the study conduct was not possible or applicable.

