## Supplementary file 1 for "Is age the most important risk factor in COVID-19 patients? The relevance of comorbidity burden: A retrospective analysis of 10,090 hospitalizations"

### Supplementary Figures: methods

#### **Figure S1.** Prevalence of SARS-CoV-2 variants of concern throughout the investigated period. A: Pango lineages. B: WHO nomenclature.

**A**
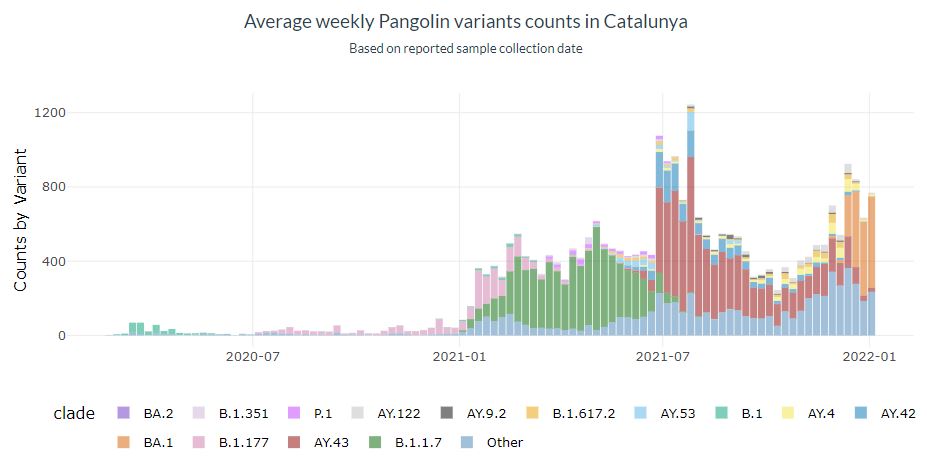

**B**

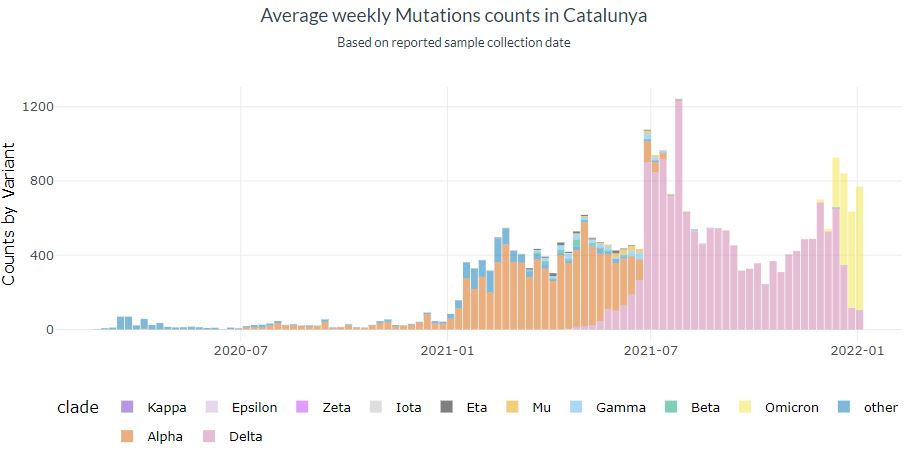

### Supplementary Figures: individual distribution across the Queralt DxS groups through the successive waves

#### **Figure S2.** Individual distribution across the Queralt DxS groups, according to age and sex. First wave.

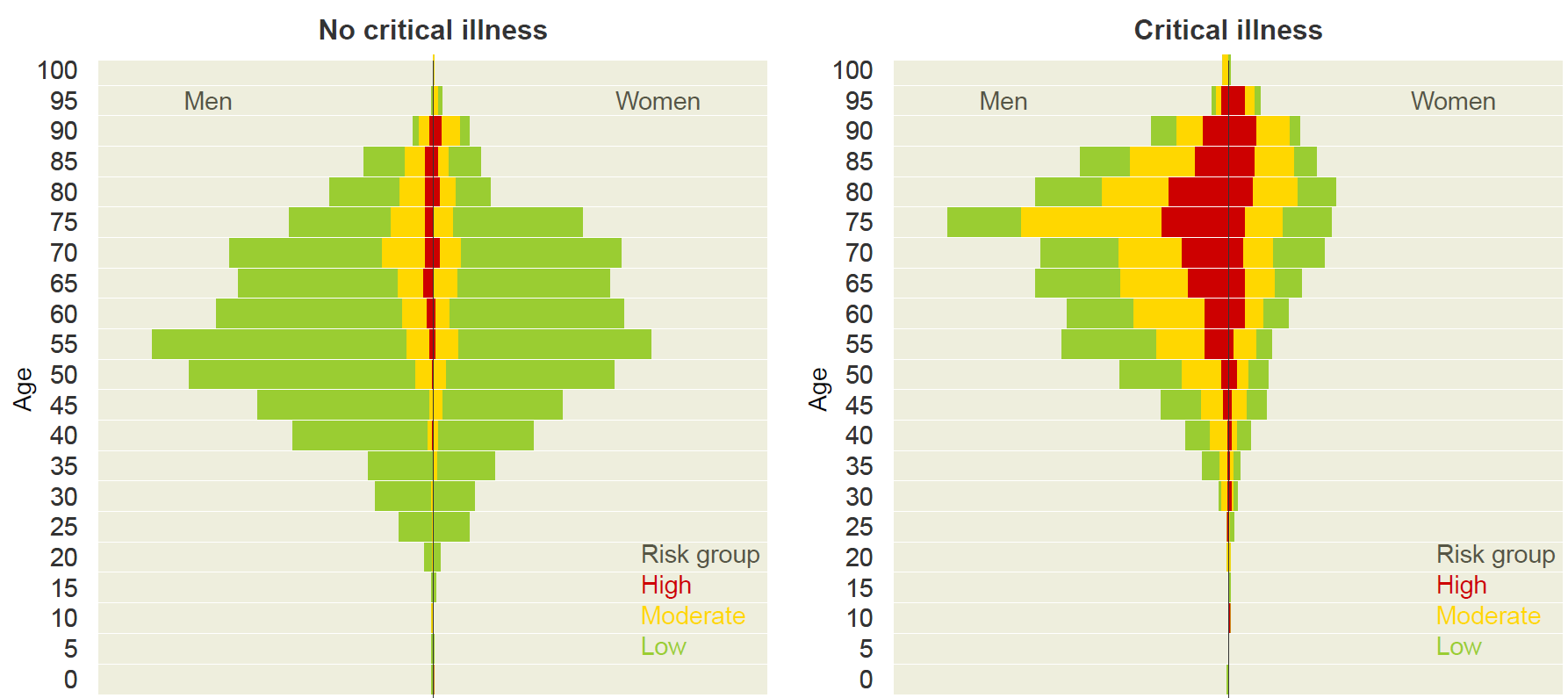

#### **Figure S3.** Individual distribution across the Queralt DxS groups, according to age and sex. Second wave.

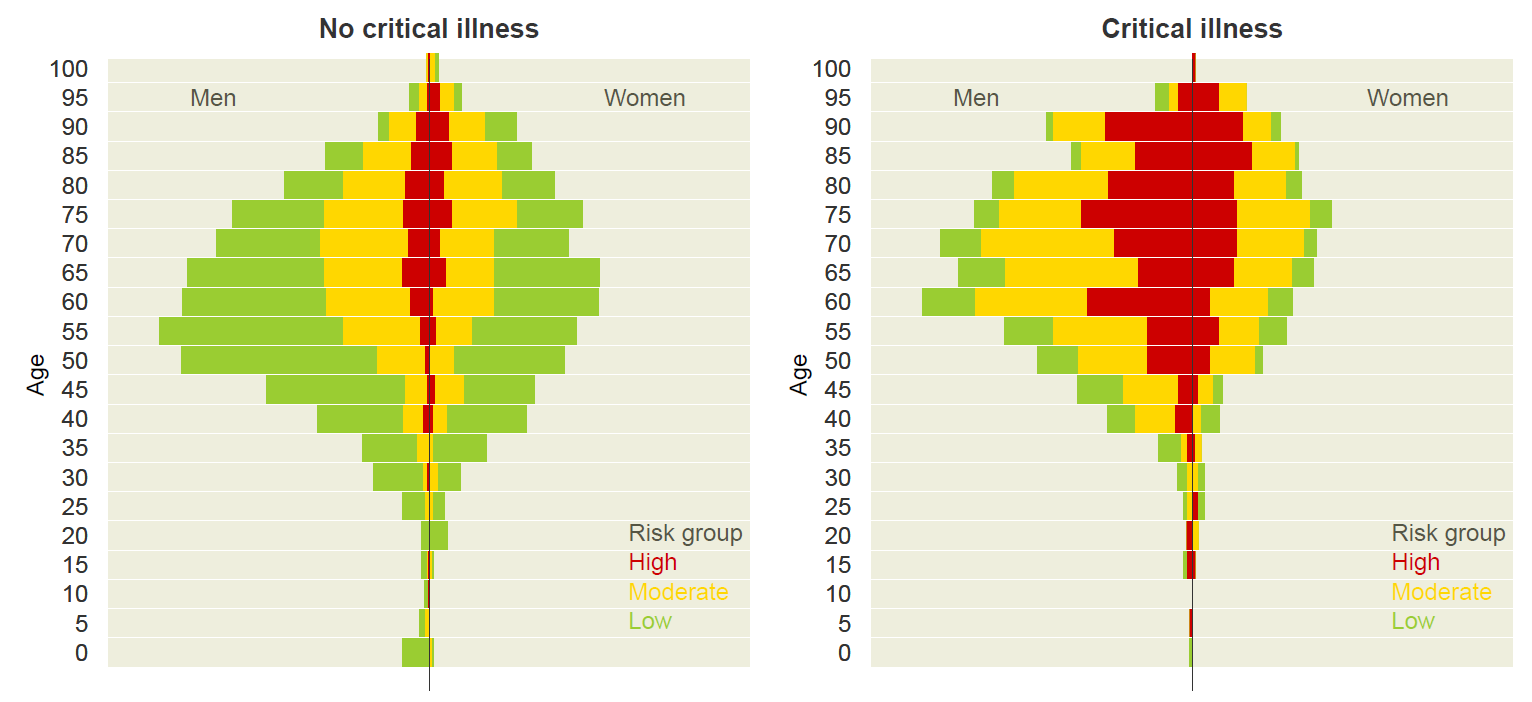

#### **Figure S4.** Individual distribution across the Queralt DxS groups, according to age and sex. Third wave.

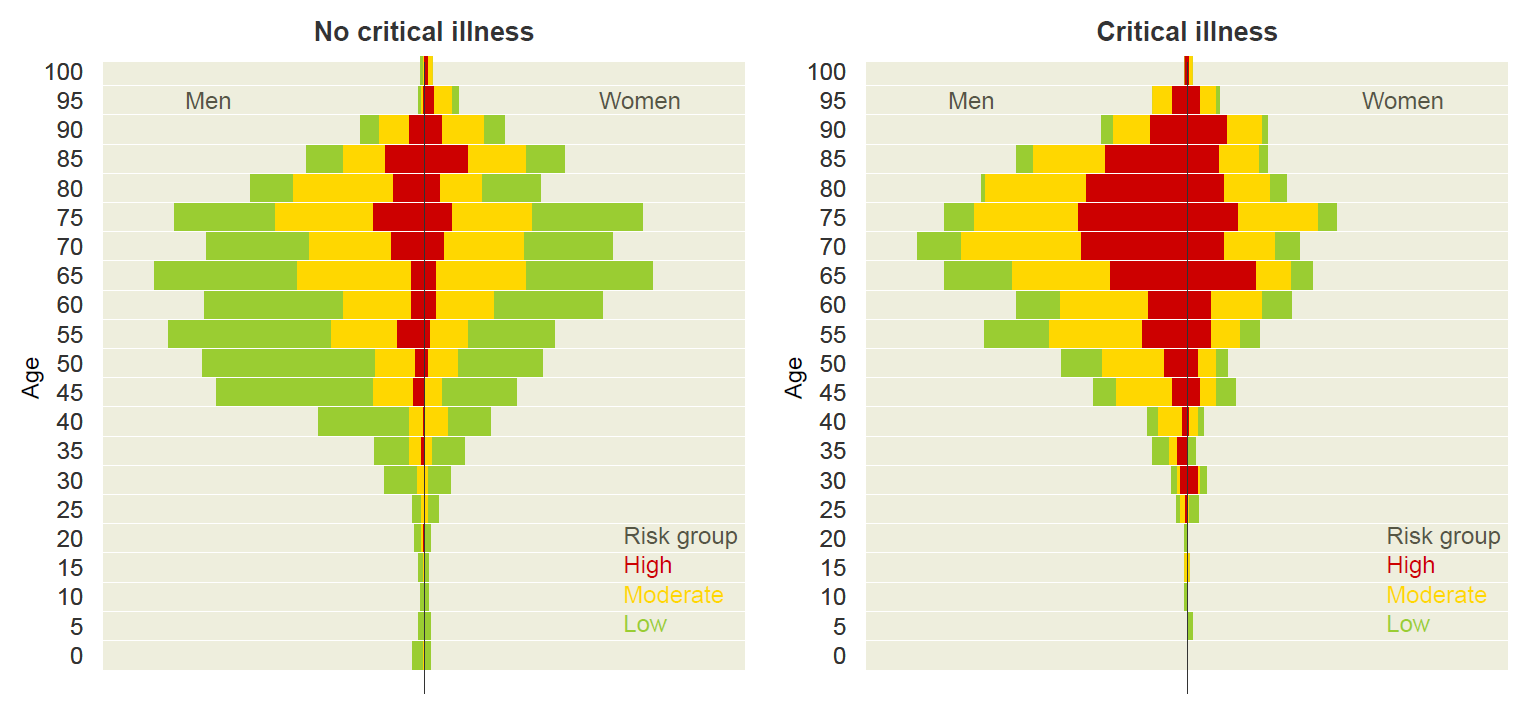

#### **Figure S5.** Individual distribution across the Queralt DxS groups, according to age and sex. Fourth wave.

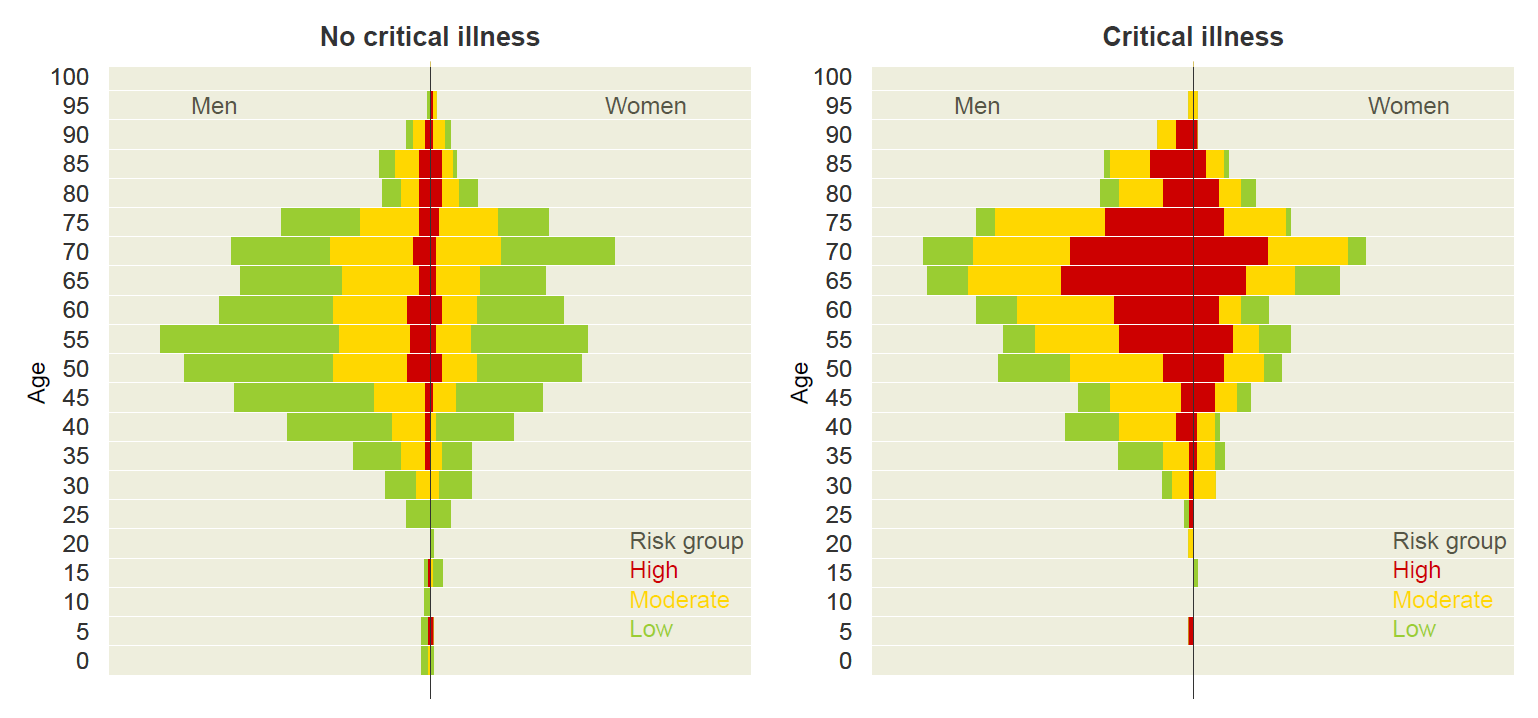

#### **Figure S6.** Individual distribution across the Queralt DxS groups, according to age and sex. Fifth wave.

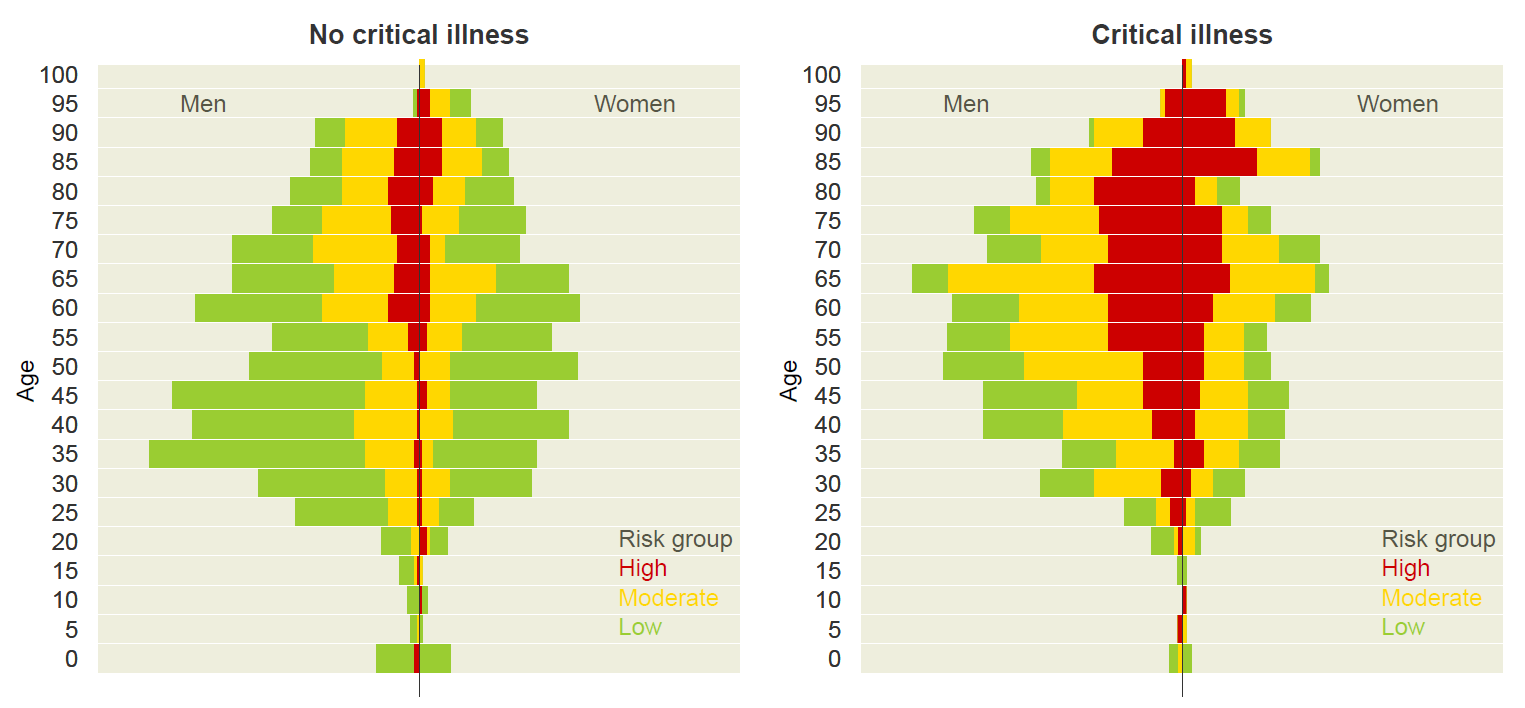

#### **Figure S7.** Individual distribution across the Queralt DxS groups, according to age and sex. Sixth wave

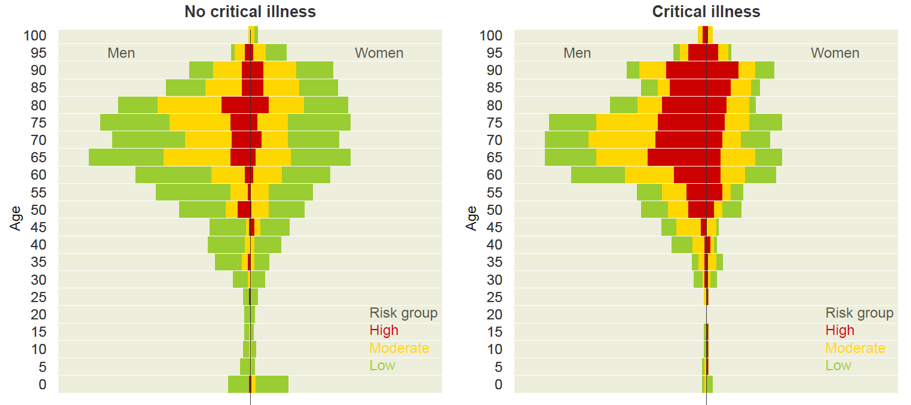

### Supplementary Figures: Unadjusted effects

#### **Figure S8.** Bivariate analysis for the three investigated outcomes and the composite of critical illness.

Dots indicate the unadjusted proportion and horizontal lines their 95% confidence intervals. Dotted vertical lines indicate the incidence of each outcome for the primary study sample.

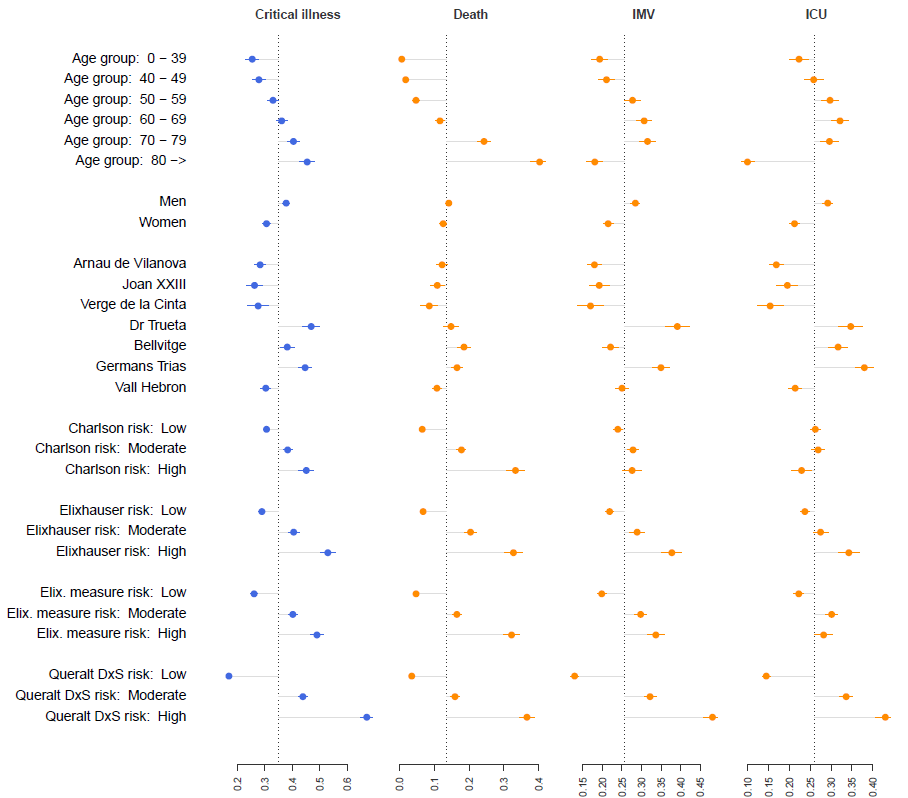

#### **Figure S9.** Bivariate analysis for the three investigated outcomes and the composite of critical illness among individuals admitted during the **first wave**.

Dots indicate the unadjusted proportion and horizontal lines their 95% confidence intervals. Dotted vertical lines indicate the incidence of each outcome for the primary study sample.

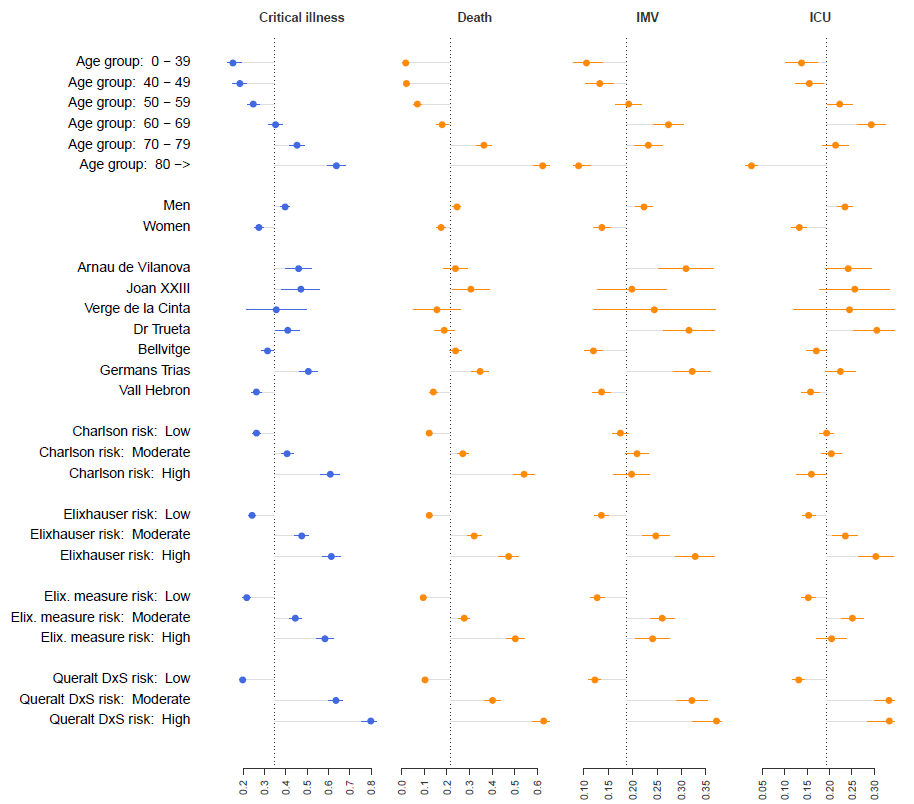

#### **Figure S10.** Bivariate analysis for the three investigated outcomes and the composite of critical illness among individuals admitted during the **second wave**.

Dots indicate the unadjusted proportion and horizontal lines their 95% confidence intervals. Dotted vertical lines indicate the incidence of each outcome for the primary study sample.

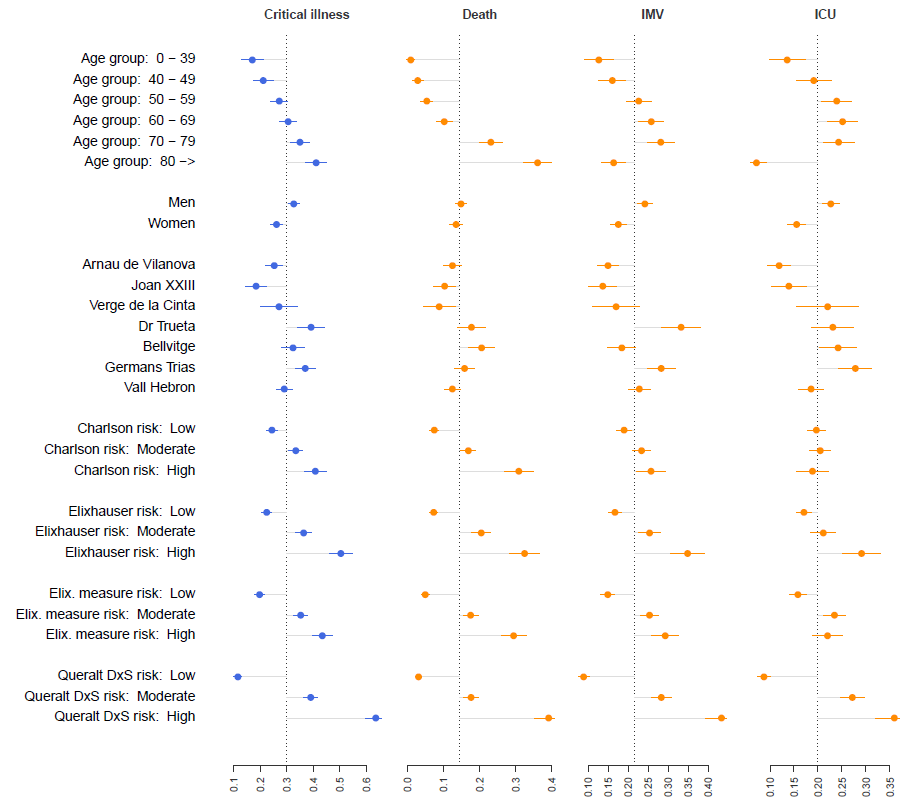

#### **Figure S11.** Bivariate analysis for the three investigated outcomes and the composite of critical illness among individuals admitted during the **third wave**.

Dots indicate the unadjusted proportion and horizontal lines their 95% confidence intervals. Dotted vertical lines indicate the incidence of each outcome for the primary study sample.

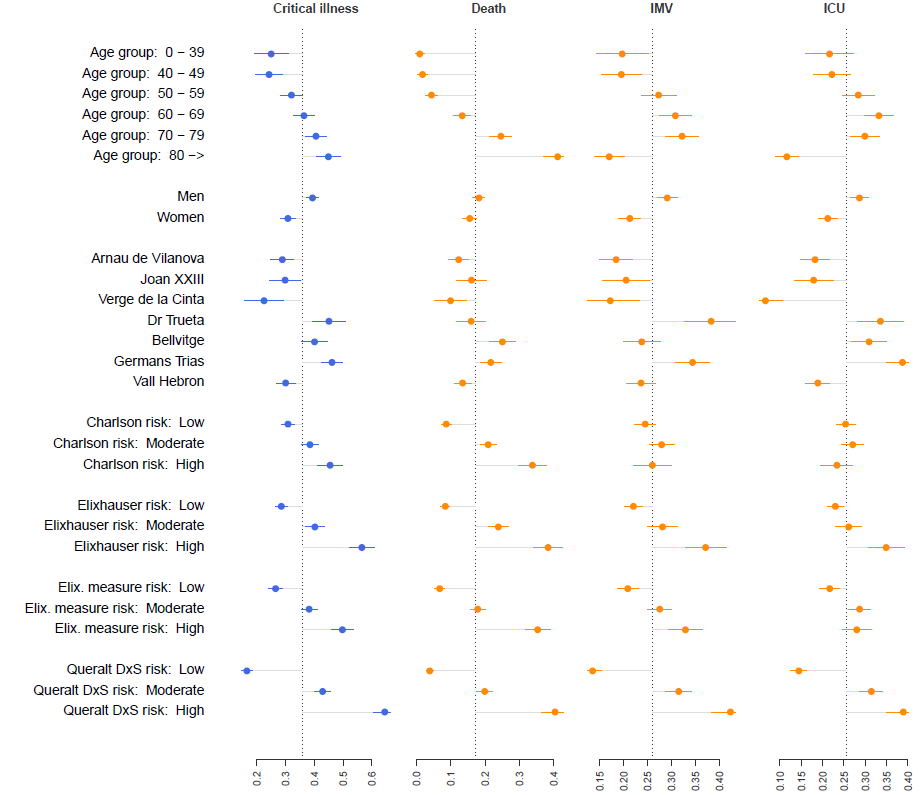

#### **Figure S12.** Bivariate analysis for the three investigated outcomes and the composite of critical illness among individuals admitted during the **fourth wave**.

Dots indicate the unadjusted proportion and horizontal lines their 95% confidence intervals. Dotted vertical lines indicate the incidence of each outcome for the primary study sample.

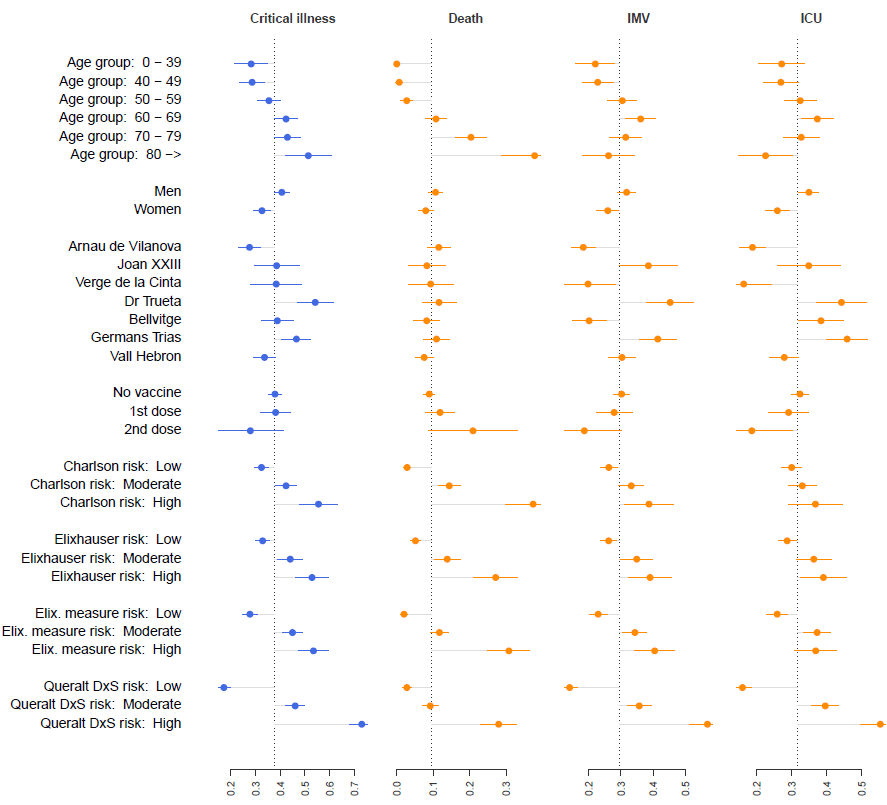

#### **Figure S13.** Bivariate analysis for the three investigated outcomes and the composite of critical illness among individuals admitted during the **fifth wave**.

Dots indicate the unadjusted proportion and horizontal lines their 95% confidence intervals. Dotted vertical lines indicate the incidence of each outcome for the primary study sample.

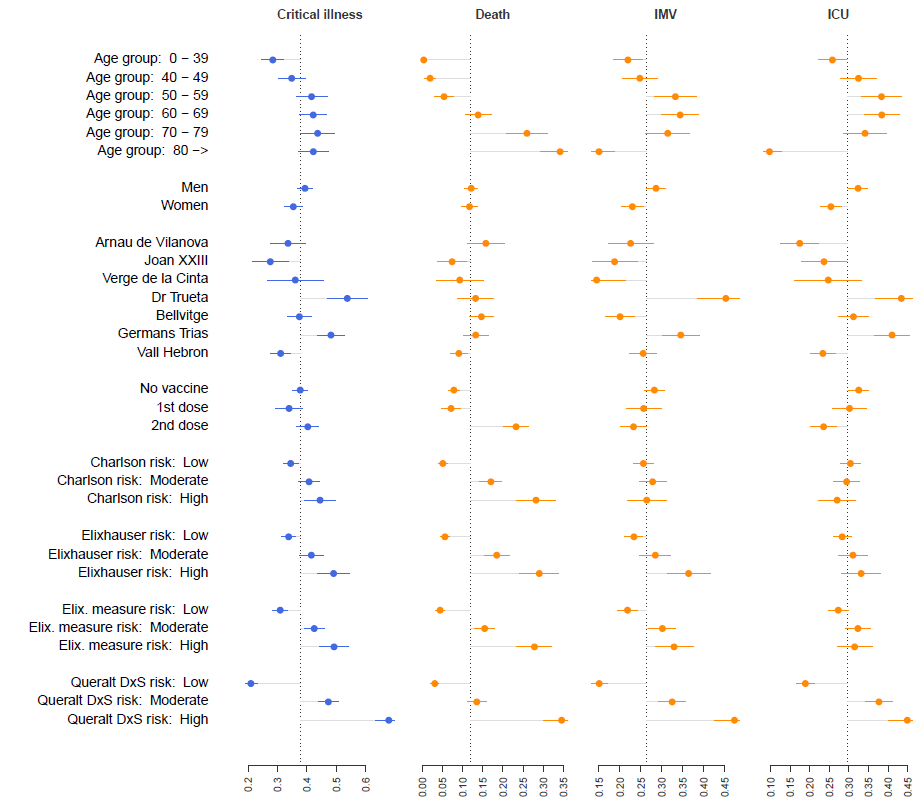

#### **Figure S14.** Bivariate analysis for the three investigated outcomes and the composite of critical illness among individuals admitted during the **sixth wave**.

Dots indicate the unadjusted proportion and horizontal lines their 95% confidence intervals. Dotted vertical lines indicate the incidence of each outcome for the primary study sample.

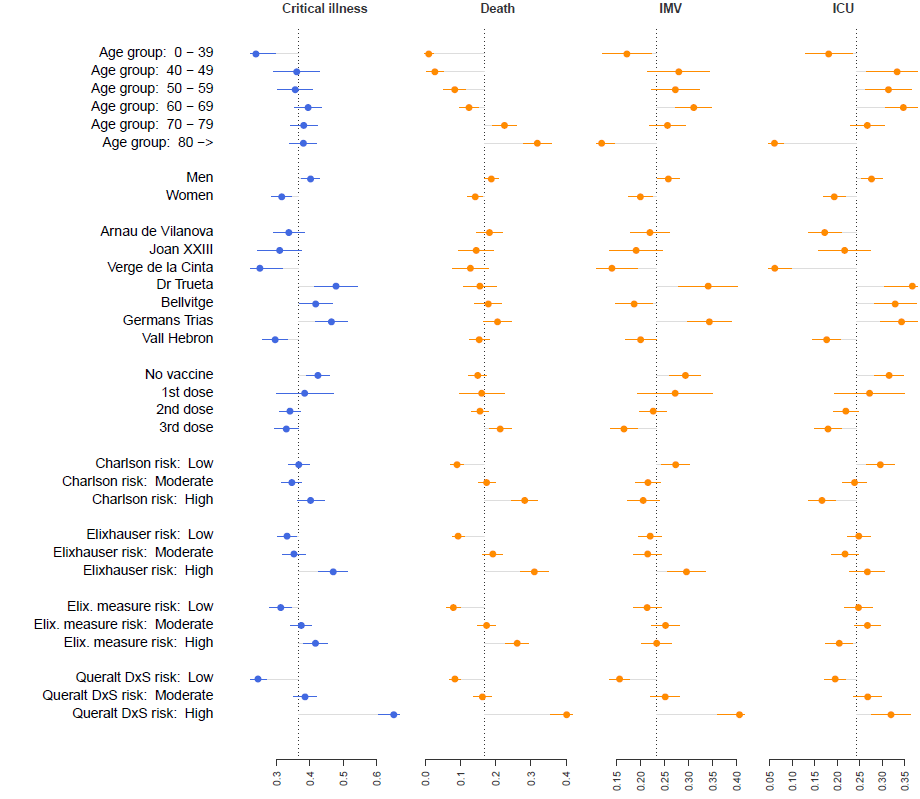

### Supplementary Figures: Adjusted standardized coefficients of age and sex

#### **Figure S15.** Effect of age (standardized coefficients for age groups) and sex on critical illness using multiple logistic regressions. **First wave**.

The baseline model included only age, sex, and hospital, whereas the other models were built by adjusting the baseline model for each of the multimorbidity measures: the Charlson index, the Elixhauser index, the unweighted count of Elixhauser diagnoses (Elixhauser measure), and the Queralt DxS.

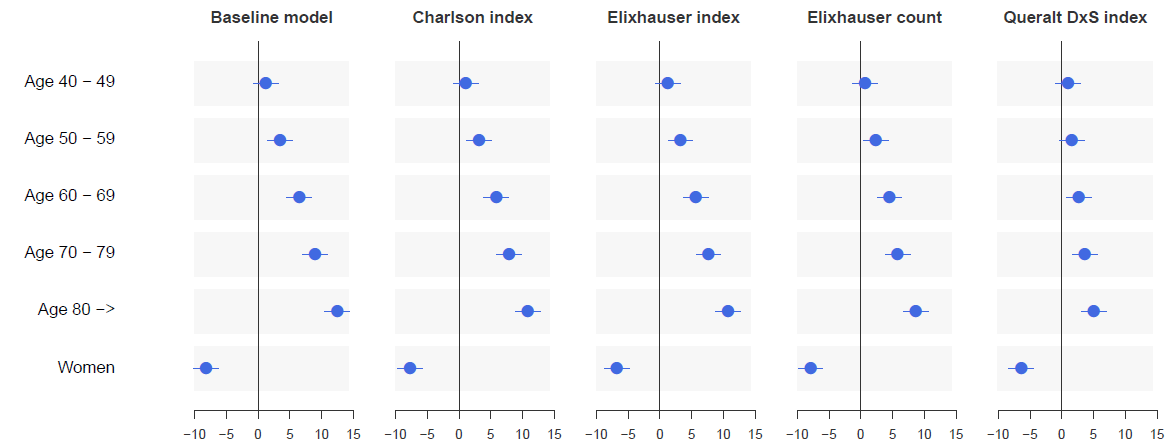

#### **Figure S16.** Effect of age (standardized coefficients for age groups) and sex on critical illness using multiple logistic regressions. **Second wave**.

The baseline model included only age, sex, and hospital, whereas the other models were built by adjusting the baseline model for each of the multimorbidity measures: the Charlson index, the Elixhauser index, the unweighted count of Elixhauser diagnoses (Elixhauser measure), and the Queralt DxS.

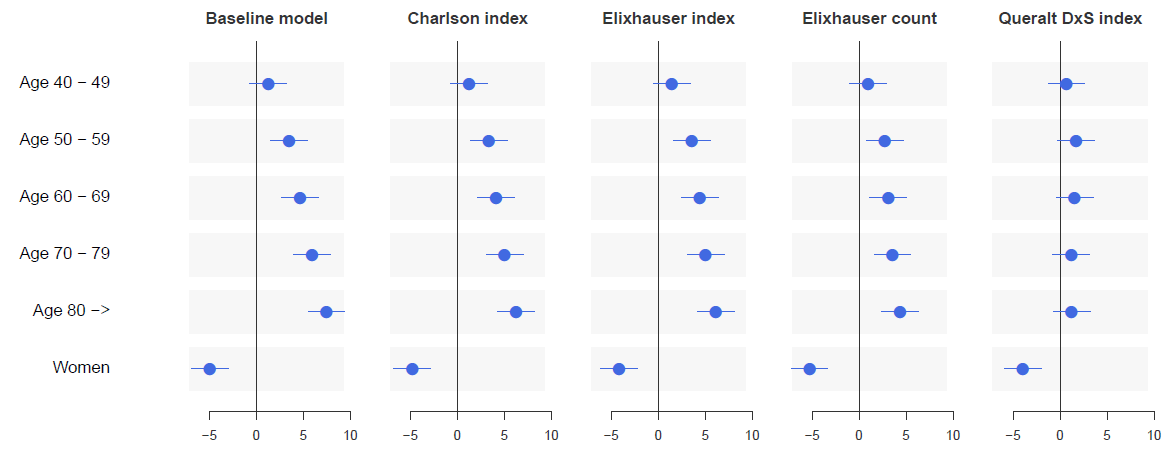

#### **Figure S17.** Effect of age (standardized coefficients for age groups) and sex on critical illness using multiple logistic regressions. **Third wave**.

The baseline model included only age, sex, and hospital, whereas the other models were built by adjusting the baseline model for each of the multimorbidity measures: the Charlson index, the Elixhauser index, the unweighted count of Elixhauser diagnoses (Elixhauser measure), and the Queralt DxS.

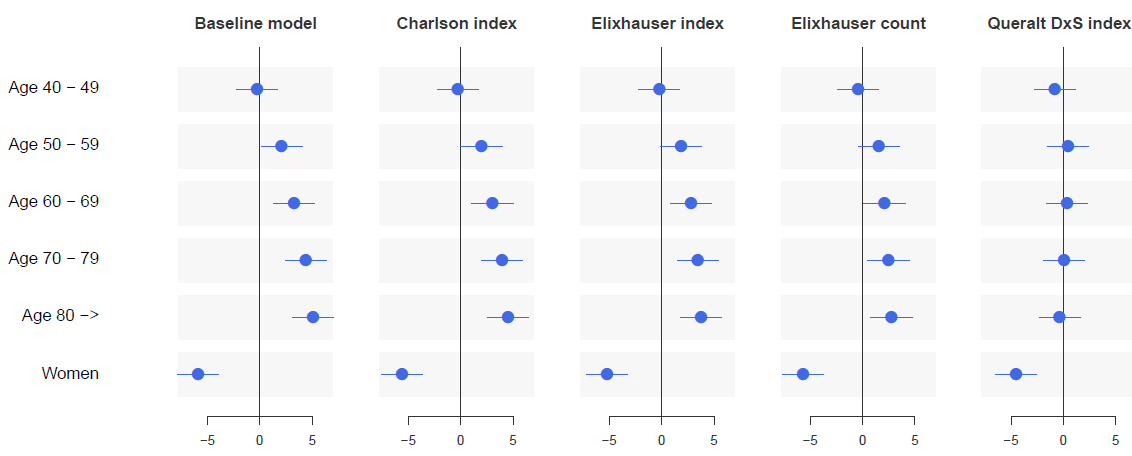

#### **Figure S18.** Effect of age (standardized coefficients for age groups) and sex on critical illness using multiple logistic regressions. **Fourth wave**.

The baseline model included only age, sex, and hospital, whereas the other models were built by adjusting the baseline model for each of the multimorbidity measures: the Charlson index, the Elixhauser index, the unweighted count of Elixhauser diagnoses (Elixhauser measure), and the Queralt DxS.

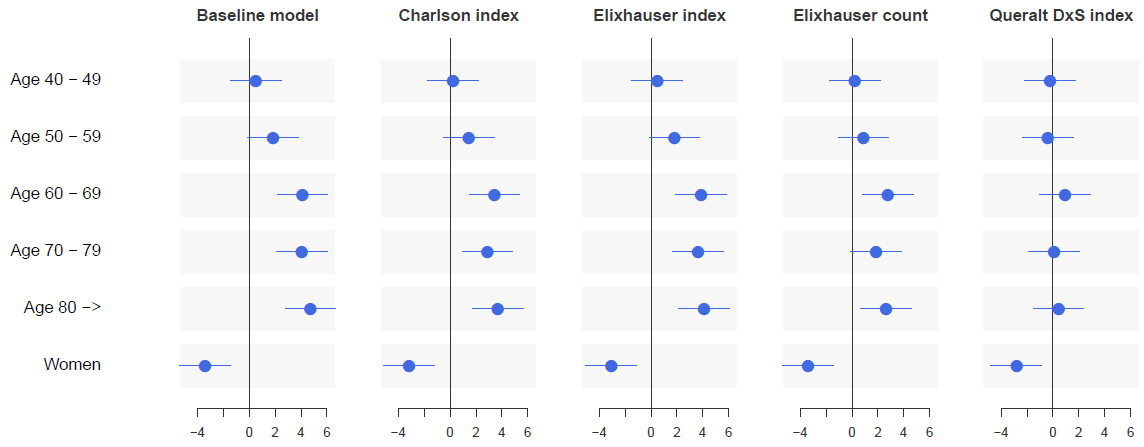

#### **Figure S19.** Effect of age (standardized coefficients for age groups) and sex on critical illness using multiple logistic regressions. **Fifth wave**.

The baseline model included only age, sex, and hospital, whereas the other models were built by adjusting the baseline model for each of the multimorbidity measures: the Charlson index, the Elixhauser index, the unweighted count of Elixhauser diagnoses (Elixhauser measure), and the Queralt DxS.

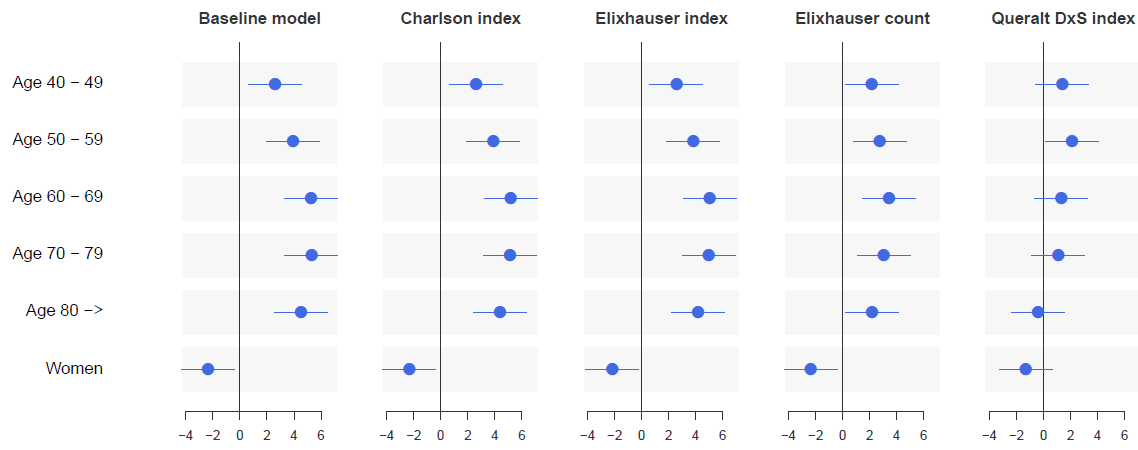

#### **Figure S20.** Effect of age (standardized coefficients for age groups) and sex on critical illness using multiple logistic regressions. **Sixth wave**.

The baseline model included only age, sex, and hospital, whereas the other models were built by adjusting the baseline model for each of the multimorbidity measures: the Charlson index, the Elixhauser index, the unweighted count of Elixhauser diagnoses (Elixhauser measure), and the Queralt DxS.

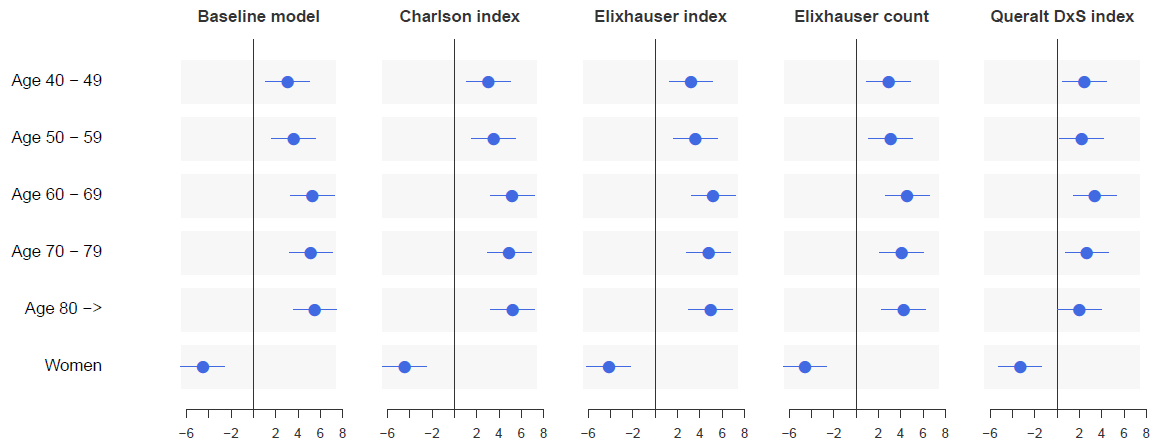

### Supplementary Tables: Main characteristics of individuals admitted in the successive waves

#### **Table S1.** Main demographic, clinical, and epidemiological characteristics of individuals admitted during the first wave

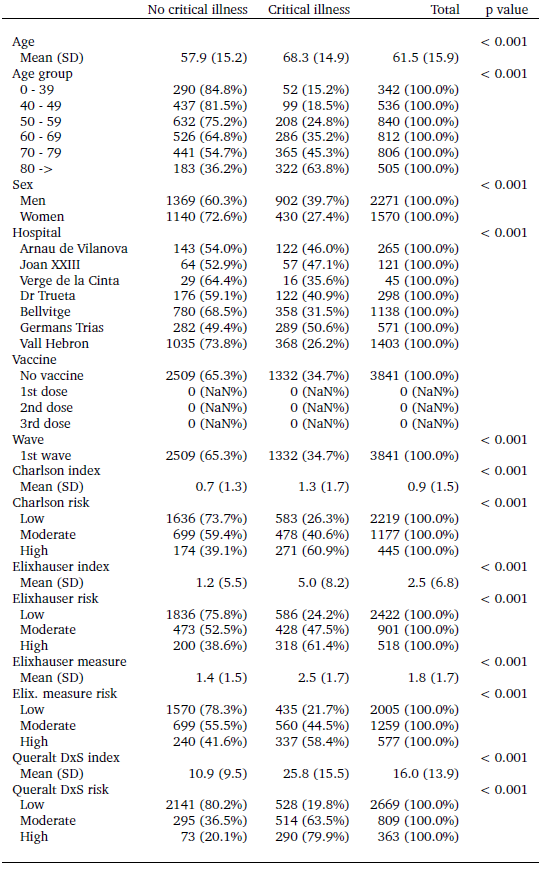

#### **Table S2.** Main demographic, clinical, and epidemiological characteristics of individuals admitted during the second wave

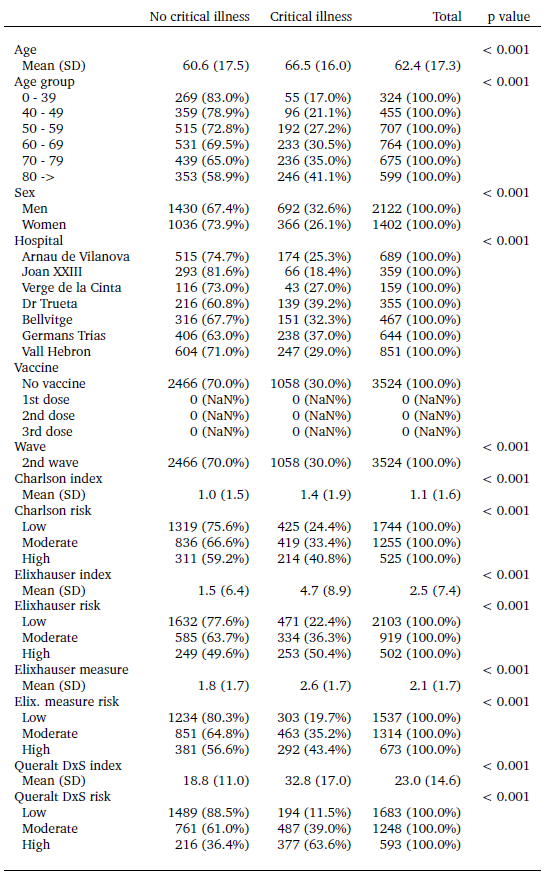

#### **Table S3.** Main demographic, clinical, and epidemiological characteristics of individuals admitted during the third wave.

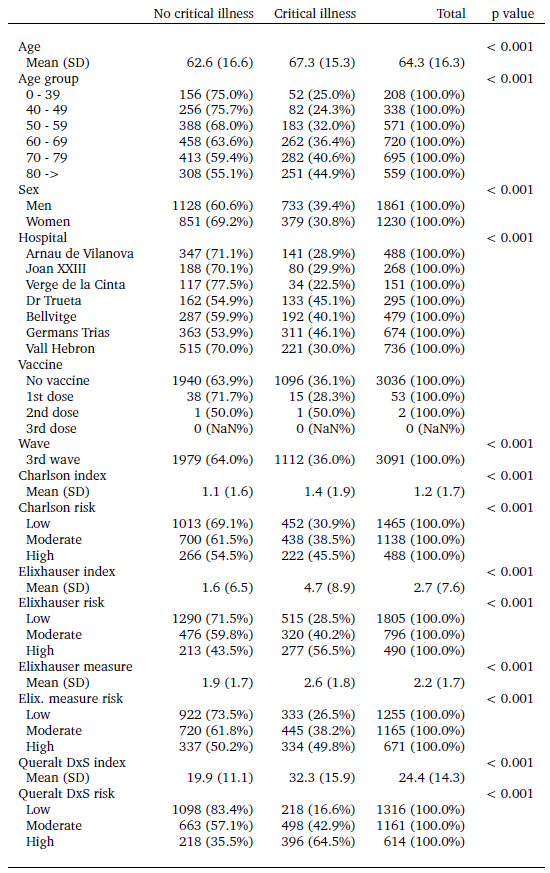

#### **Table S4.** Main demographic, clinical, and epidemiological characteristics of individuals admitted during the fourth wave.

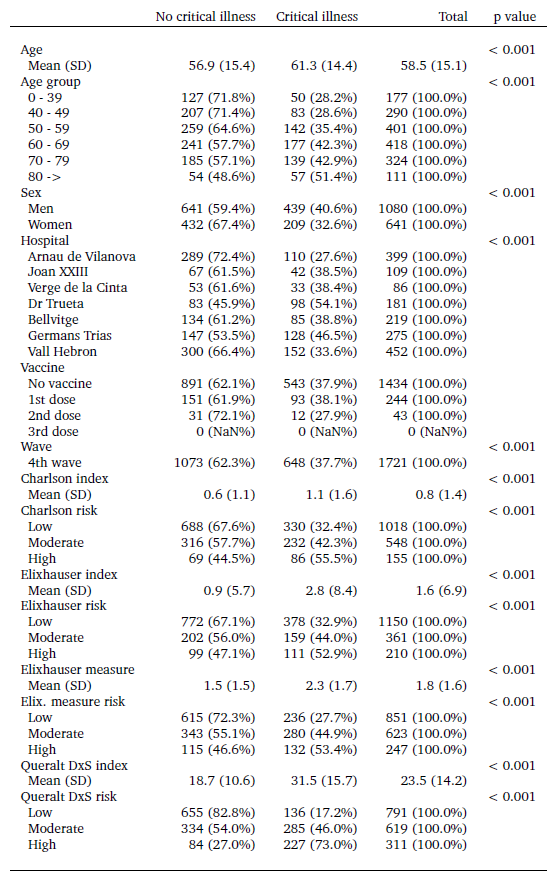

#### **Table S5.** Main demographic, clinical, and epidemiological characteristics of individuals admitted during the fifth wave

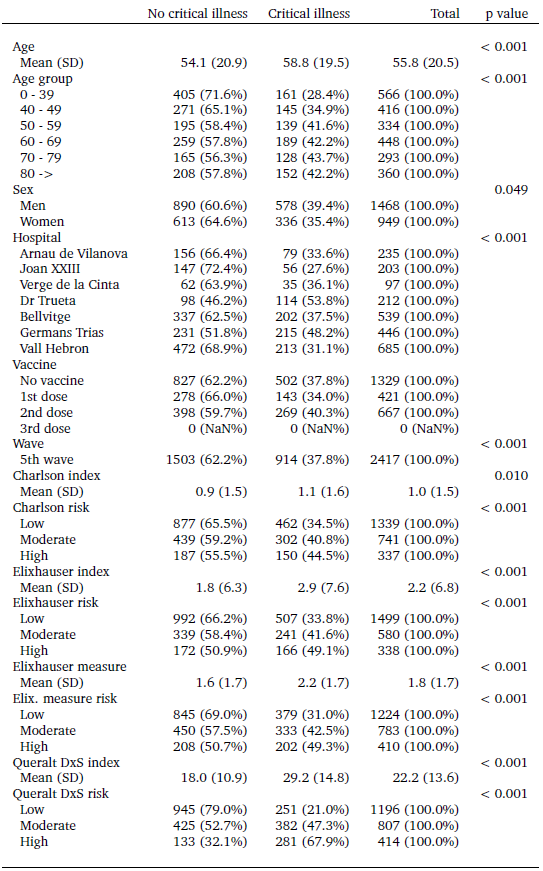

#### **Table S6.** Main demographic, clinical, and epidemiological characteristics of individuals admitted during the sixth wave.

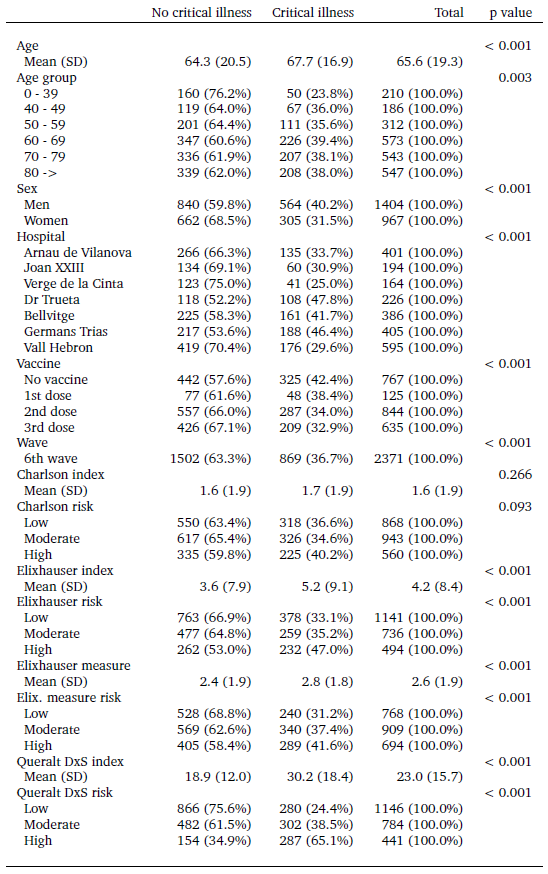

### Supplementary Tables: Performance of the models

#### **Table S7.** Performance of the models for explaining critical illness. **First wave.**

**AUPRC:** area under the precision-recall curve. **AUROC**: area under the receiving operating characteristics curve**. BIC:** bayesian criteria. **Int.:** models accounting for interactions between age and the comorbidity measure. The point estimate cells are colored based on a gradient that ranges from red (poorer performance, corresponding to lower values for the AUROCC and AUPRC, and higher values for the BIC) to green (better performance, corresponding to higher values for the AUROCC and AUPRC, and lower values for the BIC). All models have been adjusted for the hospital in which admission occurred.

|  | **BIC** |  | **AUROCC** | | **AUPRC** | |
| --- | --- | --- | --- | --- | --- | --- |
|  |  |  | Estimate | 95% CI | Estimate | 95% CI |
| Age | 4576 |  | 0,711 | 0.693 - 0.729 | 0,547 | 0.520 - 0.571 |
| Sex | 4825 |  | 0,637 | 0.620 - 0.655 | 0,465 | 0.438 - 0.490 |
| Charlson | 4731 |  | 0,67 | 0.653 - 0.689 | 0,506 | 0.479 - 0.535 |
| Elixhauser (index) | 4618 |  | 0,688 | 0.671 - 0.707 | 0,552 | 0.524 - 0.572 |
| Elixhauser (count) | 4543 |  | 0,716 | 0.698 - 0.731 | 0,55 | 0.521 - 0.578 |
| Queralt DxS | 3788 |  | 0,839 | 0.827 - 0.852 | 0,707 | 0.688 - 0.734 |
| BL model (age and sex) | 4514 |  | 0,727 | 0.711 - 0.745 | 0,562 | 0.533 - 0.591 |
| BL + Charlson | 4494 |  | 0,733 | 0.717 - 0.750 | 0,566 | 0.538 - 0.596 |
| BL + Elixhauser (index) | 4398 |  | 0,751 | 0.734 - 0.768 | 0,596 | 0.571 - 0.628 |
| BL + Elixhauser (count) | 4389 |  | 0,754 | 0.736 - 0.769 | 0,589 | 0.563 - 0.611 |
| BL + Queralt DxS | 3756 |  | 0,847 | 0.835 - 0.859 | 0,716 | 0.688 - 0.744 |
| BL + Charlson + int. | 4526 |  | 0,735 | 0.717 - 0.750 | 0,566 | 0.548 - 0.597 |
| BL + Elixhauser (index) + int. | 4412 |  | 0,751 | 0.735 - 0.767 | 0,607 | 0.575 - 0.634 |
| BL + Elixhauser (count) + int. | 4399 |  | 0,756 | 0.740 - 0.772 | 0,592 | 0.565 - 0.630 |
| BL + Queralt DxS + int. | 3731 |  | 0,847 | 0.834 - 0.859 | 0,716 | 0.690 - 0.737 |

#### **Table S8.** Performance of the models for explaining critical illness. **Second wave.**

**AUPRC:** area under the precision-recall curve. **AUROC**: area under the receiving operating characteristics curve**. BIC:** bayesian criteria. **Int.:** models accounting for interactions between age and the comorbidity measure. The point estimate cells are colored based on a gradient that ranges from red (poorer performance, corresponding to lower values for the AUROCC and AUPRC, and higher values for the BIC) to green (better performance, corresponding to higher values for the AUROCC and AUPRC, and lower values for the BIC). All models have been adjusted for the hospital in which admission occurred.

|  | **BIC** |  | **AUROCC** | | **AUPRC** | |
| --- | --- | --- | --- | --- | --- | --- |
|  |  |  | Estimate | 95% CI | Estimate | 95% CI |
| Age | 4576 |  | 0,711 | 0.693 - 0.729 | 0,547 | 0.520 - 0.571 |
| Sex | 4825 |  | 0,637 | 0.620 - 0.655 | 0,465 | 0.438 - 0.490 |
| Charlson | 4731 |  | 0,67 | 0.653 - 0.689 | 0,506 | 0.479 - 0.535 |
| Elixhauser (index) | 4618 |  | 0,688 | 0.671 - 0.707 | 0,552 | 0.524 - 0.572 |
| Elixhauser (count) | 4543 |  | 0,716 | 0.698 - 0.731 | 0,55 | 0.521 - 0.578 |
| Queralt DxS | 3788 |  | 0,839 | 0.827 - 0.852 | 0,707 | 0.688 - 0.734 |
| BL model (age and sex) | 4514 |  | 0,727 | 0.711 - 0.745 | 0,562 | 0.533 - 0.591 |
| BL + Charlson | 4494 |  | 0,733 | 0.717 - 0.750 | 0,566 | 0.538 - 0.596 |
| BL + Elixhauser (index) | 4398 |  | 0,751 | 0.734 - 0.768 | 0,596 | 0.571 - 0.628 |
| BL + Elixhauser (count) | 4389 |  | 0,754 | 0.736 - 0.769 | 0,589 | 0.563 - 0.611 |
| BL + Queralt DxS | 3756 |  | 0,847 | 0.835 - 0.859 | 0,716 | 0.688 - 0.744 |
| BL + Charlson + int. | 4526 |  | 0,735 | 0.717 - 0.750 | 0,566 | 0.548 - 0.597 |
| BL + Elixhauser (index) + int. | 4412 |  | 0,751 | 0.735 - 0.767 | 0,607 | 0.575 - 0.634 |
| BL + Elixhauser (count) + int. | 4399 |  | 0,756 | 0.740 - 0.772 | 0,592 | 0.565 - 0.630 |
| BL + Queralt DxS + int. | 3731 |  | 0,847 | 0.834 - 0.859 | 0,716 | 0.690 - 0.737 |

#### **Table S9.** Performance of the models for explaining critical illness. **Third wave.**

**AUPRC:** area under the precision-recall curve. **AUROC**: area under the receiving operating characteristics curve**. BIC:** bayesian criteria. **Int.:** models accounting for interactions between age and the comorbidity measure. The point estimate cells are colored based on a gradient that ranges from red (poorer performance, corresponding to lower values for the AUROCC and AUPRC, and higher values for the BIC) to green (better performance, corresponding to higher values for the AUROCC and AUPRC, and lower values for the BIC). All models have been adjusted for the hospital in which admission occurred.

|  | **BIC** |  | **AUROCC** | | **AUPRC** | |
| --- | --- | --- | --- | --- | --- | --- |
|  |  |  | Estimate | 95% CI | Estimate | 95% CI |
| Age | 4576 |  | 0,711 | 0.693 - 0.729 | 0,547 | 0.520 - 0.571 |
| Sex | 4825 |  | 0,637 | 0.620 - 0.655 | 0,465 | 0.438 - 0.490 |
| Charlson | 4731 |  | 0,67 | 0.653 - 0.689 | 0,506 | 0.479 - 0.535 |
| Elixhauser (index) | 4618 |  | 0,688 | 0.671 - 0.707 | 0,552 | 0.524 - 0.572 |
| Elixhauser (count) | 4543 |  | 0,716 | 0.698 - 0.731 | 0,55 | 0.521 - 0.578 |
| Queralt DxS | 3788 |  | 0,839 | 0.827 - 0.852 | 0,707 | 0.688 - 0.734 |
| BL model (age and sex) | 4514 |  | 0,727 | 0.711 - 0.745 | 0,562 | 0.533 - 0.591 |
| BL + Charlson | 4494 |  | 0,733 | 0.717 - 0.750 | 0,566 | 0.538 - 0.596 |
| BL + Elixhauser (index) | 4398 |  | 0,751 | 0.734 - 0.768 | 0,596 | 0.571 - 0.628 |
| BL + Elixhauser (count) | 4389 |  | 0,754 | 0.736 - 0.769 | 0,589 | 0.563 - 0.611 |
| BL + Queralt DxS | 3756 |  | 0,847 | 0.835 - 0.859 | 0,716 | 0.688 - 0.744 |
| BL + Charlson + int. | 4526 |  | 0,735 | 0.717 - 0.750 | 0,566 | 0.548 - 0.597 |
| BL + Elixhauser (index) + int. | 4412 |  | 0,751 | 0.735 - 0.767 | 0,607 | 0.575 - 0.634 |
| BL + Elixhauser (count) + int. | 4399 |  | 0,756 | 0.740 - 0.772 | 0,592 | 0.565 - 0.630 |
| BL + Queralt DxS + int. | 3731 |  | 0,847 | 0.834 - 0.859 | 0,716 | 0.690 - 0.737 |

#### **Table S10.** Performance of the models for explaining critical illness. **Fourth wave.**

**AUPRC:** area under the precision-recall curve. **AUROC**: area under the receiving operating characteristics curve**. BIC:** bayesian criteria. **Int.:** models accounting for interactions between age and the comorbidity measure. The point estimate cells are colored based on a gradient that ranges from red (poorer performance, corresponding to lower values for the AUROCC and AUPRC, and higher values for the BIC) to green (better performance, corresponding to higher values for the AUROCC and AUPRC, and lower values for the BIC). All models have been adjusted for the hospital in which admission occurred.

|  | **BIC** |  | **AUROCC** | | **AUPRC** | |
| --- | --- | --- | --- | --- | --- | --- |
|  |  |  | Estimate | 95% CI | Estimate | 95% CI |
| Age | 4576 |  | 0,711 | 0.693 - 0.729 | 0,547 | 0.520 - 0.571 |
| Sex | 4825 |  | 0,637 | 0.620 - 0.655 | 0,465 | 0.438 - 0.490 |
| Charlson | 4731 |  | 0,67 | 0.653 - 0.689 | 0,506 | 0.479 - 0.535 |
| Elixhauser (index) | 4618 |  | 0,688 | 0.671 - 0.707 | 0,552 | 0.524 - 0.572 |
| Elixhauser (count) | 4543 |  | 0,716 | 0.698 - 0.731 | 0,55 | 0.521 - 0.578 |
| Queralt DxS | 3788 |  | 0,839 | 0.827 - 0.852 | 0,707 | 0.688 - 0.734 |
| BL model (age and sex) | 4514 |  | 0,727 | 0.711 - 0.745 | 0,562 | 0.533 - 0.591 |
| BL + Charlson | 4494 |  | 0,733 | 0.717 - 0.750 | 0,566 | 0.538 - 0.596 |
| BL + Elixhauser (index) | 4398 |  | 0,751 | 0.734 - 0.768 | 0,596 | 0.571 - 0.628 |
| BL + Elixhauser (count) | 4389 |  | 0,754 | 0.736 - 0.769 | 0,589 | 0.563 - 0.611 |
| BL + Queralt DxS | 3756 |  | 0,847 | 0.835 - 0.859 | 0,716 | 0.688 - 0.744 |
| BL + Charlson + int. | 4526 |  | 0,735 | 0.717 - 0.750 | 0,566 | 0.548 - 0.597 |
| BL + Elixhauser (index) + int. | 4412 |  | 0,751 | 0.735 - 0.767 | 0,607 | 0.575 - 0.634 |
| BL + Elixhauser (count) + int. | 4399 |  | 0,756 | 0.740 - 0.772 | 0,592 | 0.565 - 0.630 |
| BL + Queralt DxS + int. | 3731 |  | 0,847 | 0.834 - 0.859 | 0,716 | 0.690 - 0.737 |

#### **Table S11.** Performance of the models for explaining critical illness. **Fifth wave.**

**AUPRC:** area under the precision-recall curve. **AUROC**: area under the receiving operating characteristics curve**. BIC:** bayesian criteria. **Int.:** models accounting for interactions between age and the comorbidity measure. The point estimate cells are colored based on a gradient that ranges from red (poorer performance, corresponding to lower values for the AUROCC and AUPRC, and higher values for the BIC) to green (better performance, corresponding to higher values for the AUROCC and AUPRC, and lower values for the BIC). All models have been adjusted for the hospital in which admission occurred.

|  | **BIC** |  | **AUROCC** | | **AUPRC** | |
| --- | --- | --- | --- | --- | --- | --- |
|  |  |  | Estimate | 95% CI | Estimate | 95% CI |
| Age | 4576 |  | 0,711 | 0.693 - 0.729 | 0,547 | 0.520 - 0.571 |
| Sex | 4825 |  | 0,637 | 0.620 - 0.655 | 0,465 | 0.438 - 0.490 |
| Charlson | 4731 |  | 0,67 | 0.653 - 0.689 | 0,506 | 0.479 - 0.535 |
| Elixhauser (index) | 4618 |  | 0,688 | 0.671 - 0.707 | 0,552 | 0.524 - 0.572 |
| Elixhauser (count) | 4543 |  | 0,716 | 0.698 - 0.731 | 0,55 | 0.521 - 0.578 |
| Queralt DxS | 3788 |  | 0,839 | 0.827 - 0.852 | 0,707 | 0.688 - 0.734 |
| BL model (age and sex) | 4514 |  | 0,727 | 0.711 - 0.745 | 0,562 | 0.533 - 0.591 |
| BL + Charlson | 4494 |  | 0,733 | 0.717 - 0.750 | 0,566 | 0.538 - 0.596 |
| BL + Elixhauser (index) | 4398 |  | 0,751 | 0.734 - 0.768 | 0,596 | 0.571 - 0.628 |
| BL + Elixhauser (count) | 4389 |  | 0,754 | 0.736 - 0.769 | 0,589 | 0.563 - 0.611 |
| BL + Queralt DxS | 3756 |  | 0,847 | 0.835 - 0.859 | 0,716 | 0.688 - 0.744 |
| BL + Charlson + int. | 4526 |  | 0,735 | 0.717 - 0.750 | 0,566 | 0.548 - 0.597 |
| BL + Elixhauser (index) + int. | 4412 |  | 0,751 | 0.735 - 0.767 | 0,607 | 0.575 - 0.634 |
| BL + Elixhauser (count) + int. | 4399 |  | 0,756 | 0.740 - 0.772 | 0,592 | 0.565 - 0.630 |
| BL + Queralt DxS + int. | 3731 |  | 0,847 | 0.834 - 0.859 | 0,716 | 0.690 - 0.737 |

#### **Table S12.** Performance of the models for explaining critical illness. **Sixth wave.**

**AUPRC:** area under the precision-recall curve. **AUROC**: area under the receiving operating characteristics curve**. BIC:** bayesian criteria. **Int.:** models accounting for interactions between age and the comorbidity measure. The point estimate cells are colored based on a gradient that ranges from red (poorer performance, corresponding to lower values for the AUROCC and AUPRC, and higher values for the BIC) to green (better performance, corresponding to higher values for the AUROCC and AUPRC, and lower values for the BIC). All models have been adjusted for the hospital in which admission occurred.

|  | **BIC** |  | **AUROCC** | | **AUPRC** | |
| --- | --- | --- | --- | --- | --- | --- |
|  |  |  | Estimate | 95% CI | Estimate | 95% CI |
| Age | 4576 |  | 0,711 | 0.693 - 0.729 | 0,547 | 0.520 - 0.571 |
| Sex | 4825 |  | 0,637 | 0.620 - 0.655 | 0,465 | 0.438 - 0.490 |
| Charlson | 4731 |  | 0,67 | 0.653 - 0.689 | 0,506 | 0.479 - 0.535 |
| Elixhauser (index) | 4618 |  | 0,688 | 0.671 - 0.707 | 0,552 | 0.524 - 0.572 |
| Elixhauser (count) | 4543 |  | 0,716 | 0.698 - 0.731 | 0,55 | 0.521 - 0.578 |
| Queralt DxS | 3788 |  | 0,839 | 0.827 - 0.852 | 0,707 | 0.688 - 0.734 |
| BL model (age and sex) | 4514 |  | 0,727 | 0.711 - 0.745 | 0,562 | 0.533 - 0.591 |
| BL + Charlson | 4494 |  | 0,733 | 0.717 - 0.750 | 0,566 | 0.538 - 0.596 |
| BL + Elixhauser (index) | 4398 |  | 0,751 | 0.734 - 0.768 | 0,596 | 0.571 - 0.628 |
| BL + Elixhauser (count) | 4389 |  | 0,754 | 0.736 - 0.769 | 0,589 | 0.563 - 0.611 |
| BL + Queralt DxS | 3756 |  | 0,847 | 0.835 - 0.859 | 0,716 | 0.688 - 0.744 |
| BL + Charlson + int. | 4526 |  | 0,735 | 0.717 - 0.750 | 0,566 | 0.548 - 0.597 |
| BL + Elixhauser (index) + int. | 4412 |  | 0,751 | 0.735 - 0.767 | 0,607 | 0.575 - 0.634 |
| BL + Elixhauser (count) + int. | 4399 |  | 0,756 | 0.740 - 0.772 | 0,592 | 0.565 - 0.630 |
| BL + Queralt DxS + int. | 3731 |  | 0,847 | 0.834 - 0.859 | 0,716 | 0.690 - 0.737 |

### Supplementary Tables: Causal mediation analysis by wave

#### **Table S13.** Causal mediation effect, direct effect, and proportion of mediation effect by comorbidity measures. **First wave**.

The proportion mediated shows the contribution of the comorbidity-mediating pathway to critical illness, estimated as the proportion between the ACME and the total effect. **ACME**: average causal mediation effect of comorbidity (mediator). **ADE**: average direct effect of age.

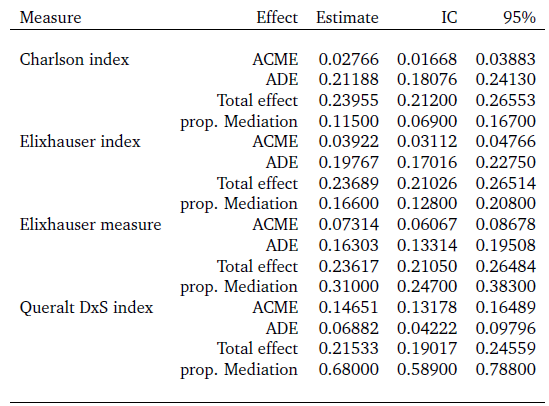

#### **Table S14.** Causal mediation effect, direct effect, and proportion of mediation effect by comorbidity measures. **Second wave**.

The proportion mediated shows the contribution of the comorbidity-mediating pathway to critical illness, estimated as the proportion between the ACME and the total effect. **ACME**: average causal mediation effect of comorbidity (mediator). **ADE**: average direct effect of age.

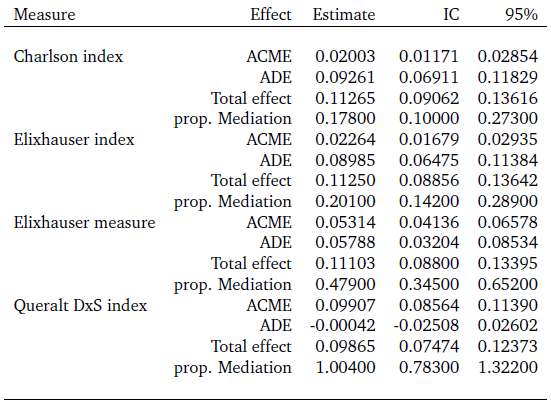

#### **Table S15.** Causal mediation effect, direct effect, and proportion of mediation effect by comorbidity measures. **Third wave**.

The proportion mediated shows the contribution of the comorbidity-mediating pathway to critical illness, estimated as the proportion between the ACME and the total effect. **ACME**: average causal mediation effect of comorbidity (mediator). **ADE**: average direct effect of age.

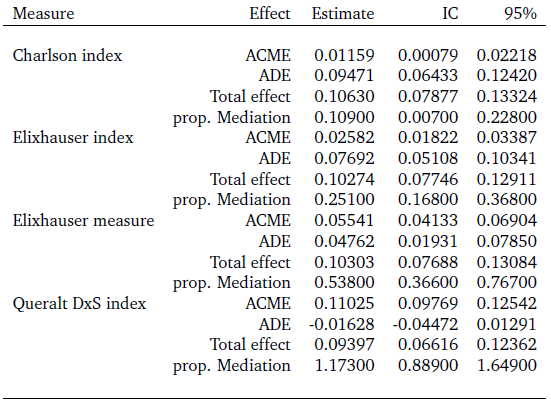

#### **Table S16.** Causal mediation effect, direct effect, and proportion of mediation effect by comorbidity measures. **Fourth wave**.

The proportion mediated shows the contribution of the comorbidity-mediating pathway to critical illness, estimated as the proportion between the ACME and the total effect. **ACME**: average causal mediation effect of comorbidity (mediator). **ADE**: average direct effect of age.

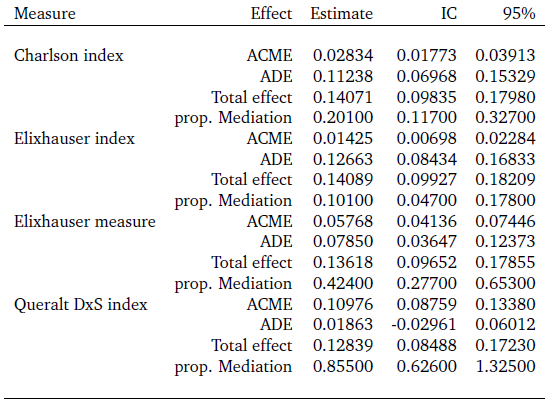

#### **Table S17.** Causal mediation effect, direct effect, and proportion of mediation effect by comorbidity measures. **Fifth wave**.

The proportion mediated shows the contribution of the comorbidity-mediating pathway to critical illness, estimated as the proportion between the ACME and the total effect. **ACME**: average causal mediation effect of comorbidity (mediator). **ADE**: average direct effect of age

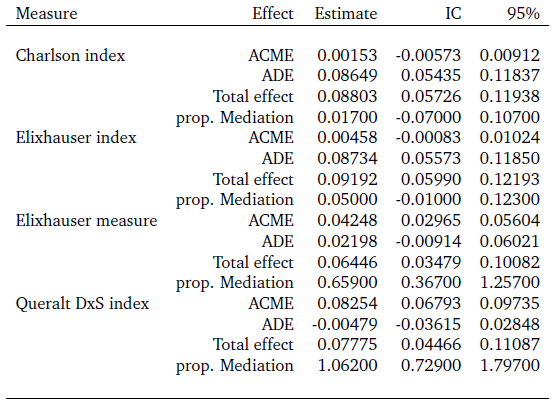

#### **Table S18.** Causal mediation effect, direct effect, and proportion of mediation effect by comorbidity measures. **Sixth wave**.

The proportion mediated shows the contribution of the comorbidity-mediating pathway to critical illness, estimated as the proportion between the ACME and the total effect. **ACME**: average causal mediation effect of comorbidity (mediator). **ADE**: average direct effect of age.

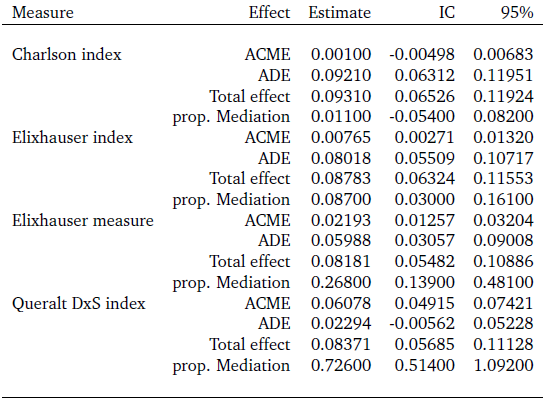
